# Differences in COVID-19 vaccination coverage by occupation in England: a national linked data study

**DOI:** 10.1101/2021.11.10.21266124

**Authors:** Vahé Nafilyan, Ted Dolby, Katie Finning, Jasper Morgan, Rhiannon Edge, Myer Glickman, Neil Pearce, Martie van Tongeren

## Abstract

**Background:** Monitoring differences in COVID-19 vaccination uptake in different groups is crucial to help inform the policy response to the pandemic. A key gap is the absence of data on uptake by occupation.

**Methods:** Using nationwide population-level data, we calculated the proportion of people who had received two doses of a COVID-19 vaccine (assessed on 31 August 2021) by detailed occupational categories in adults aged 40-64 and estimated adjusted odds ratios to examine whether these differences were driven by occupation or other factors, such as education. We also examined whether vaccination rates differed by ability to work from home.

**Results:** Our study population included 14,298,147 adults 40-64. Vaccination rates differed markedly by occupation, being higher in administrative and secretarial occupations (90.8%); professional occupations (90.7%); and managers, directors and senior officials (90.6%); and lowest (83.1%) in people working in elementary occupations. We found substantial differences in vaccination rates looking at finer occupational groups even after adjusting for confounding factors, such as education. Vaccination rates were higher in occupations which can be done from home and lower in those which cannot. Many occupations with low vaccination rates also involved contact with the public or with vulnerable people

**Conclusions:** Increasing vaccination coverage in occupations with low vaccination rates is crucial to help protecting the public and control infection, especially in occupations that cannot be done from home and involve contacts with the public. Policies such as ‘work from home if you can’ may only have limited future impact on hospitalisations and deaths

**What is already known on this subject?:** Whilst several studies highlight differences in vaccination coverage by ethnicity, religion, socio-demographic factors and certain underlying health conditions, there is very little evidence on how vaccination coverage varies by occupation, in the UK and elsewhere. The few study looking at occupational differences in vaccine hesitancy focus on healthcare workers or only examined broad occupational groups. There is currently no large-scale study on occupational differences in COVID-19 vaccination coverage in the UK.

**What this study adds?:** Using population-level linked data combining the 2011 Census, primary care records, mortality and vaccination data, we found that the vaccination rates of adults aged 40 to 64 years in England differed markedly by occupation. Vaccination rates were high in administrative and secretarial occupations, professional occupations and managers, directors and senior officials and low in people working in elementary occupations. Adjusting for other factors likely to be linked to occupation and vaccination, such as education, did not substantially alter the results. Vaccination rates were also associated with the ability to work from home, with the vaccination rate being higher in occupations which can be done performed from home. Policies aiming to increase vaccination rates in occupations that cannot be done from home and involve contacts with the public should be priorities

## Introduction

The UK began an ambitious vaccination programme to combat the COVID-19 pandemic on 8 December 2020; by 31 August 2021, 78.9% of the UK adult population had received two doses [1].

Monitoring differences in vaccine uptake between population groups is crucial to inform the policy response. Evidence suggests that rates of COVID-19 vaccination in England differ by ethnicity, religion, socio-demographic factors and certain underlying health conditions [2, 3, 4]. However, a key evidence gap is the absence of data on uptake by occupation. Producing vaccination rates by detailed occupational categories is challenging as routinely collected data used for the analysis of vaccination uptake do not contain information on occupation. The use of surveys is limited as a large sample size is needed to precisely estimate vaccination rates in small groups.

As there is evidence that people working in occupations involving contact with patients or the public are at greater risk of COVID-19 infection and death [5, 6, 7], it is important to ensure that vaccination coverage is high in these occupations. Measuring vaccination uptake by occupation also has implications for the management of the pandemic, especially as governments are starting to open up the economy, by ending the furlough scheme and encouraging workers to go back to the office. If vaccination rates are high in people working in occupations in which working from home is an option, then having workers back to the office would be expected to have less impact on cases and hospitalisation than if the vaccination rates are low. Similarly, imposing working from home where possible would be less effective if vaccination coverage is high in people who can work from home but low in those who cannot.

This study investigates differences in vaccination rates by occupation in England, using nationwide population-level data. We calculated vaccination rates by detailed occupational categories in adults aged 40 to 64 and estimated adjusted odds ratios to examine whether these differences were driven by potential confounding factors. We also examined whether vaccination rates differed by the ability to work from home and by risk of exposure to infectious diseases.

## Methods

### Data

We linked vaccination data from NHS England’s National Immunisation Management System (NIMS) to the Office for National Statistics (ONS) Public Health Data Asset (PHDA) based on NHS number. The ONS PHDA is a linked dataset combining the 2011 Census, mortality records, the General Practice Extraction Service (GPES) data for pandemic planning and research, and the Hospital Episode Statistics (HES). To obtain NHS numbers for the 2011 Census, we linked the 2011 Census to the 2011-2013 NHS Patient Registers using deterministic and probabilistic matching, with an overall linkage rate of 94.6%. All subsequent linkages were performed based on NHS numbers.

The study population consisted of adults aged 40 to 64 years, alive on 31 August 2021, who were resident in England, registered with a general practitioner, enumerated and having reported an occupation at the 2011 Census. Of 15,959,777 adults aged 40 to 64 years who received two doses of a COVID-19 vaccine in NIMS, 13,029,828 (81.6%) were linked to the ONS PHDA. We restricted our sample to individuals who were aged 40-64 years because they were likely to be in stable employment both in 2011 and 2020 [7]. Of the17,501,132 people enumerated at the 2011 Census in England and Wales, who would be aged 40-64 on 31 August 2020, we excluded 717,667 people (4.1%) who could not be linked deterministically or probabilistically to the NHS Patient register, and. 362,050 individuals (2.2%) who had died between the Census and 31 August 2021. An additional 2,121,216 people (12.6%) were not linked to the English primary care records, either because they did not live in England in 2019 (the Census included people living in England and Wales), or because they were not registered with the NHS (see sample flow diagram in Supplementary Table S1). After excluding 2,052 individuals who had received their vaccine before the beginning of the vaccination campaign (8 December 2020), our sample consisted of 14,298,147 individuals.

### Outcome

The primary outcome was being fully vaccinated against COVID-19’, that is having received two doses of a COVID-19 vaccine as recorded in the NIMS data by 31 August 2021. The vaccine roll-out started to include all aged 40 to 44 on 30^th^ April. Therefore, by the end of August 2021, we would expect most people who wanted to be vaccinated to have received their second dose. To account for the delay in receiving the first dose, and therefore not having had time to receive the second dose by then end of the study period, we also examined having received no COVID-19 vaccine and having received only the first does of the vaccine as secondary outcomes.

### Exposure

The main exposure was occupation at the time of the 2011 Census. Occupations are coded using a hierarchical classification, the Standard Occupation Classification (SOC) 2010 (7). The most detailed classification (Unit group, with 4-digit codes) includes 370 categories, whilst the most aggregated (Major group, with 1-digit codes) has only nine groups. We used both SOC major groups and unit groups in this paper.

### Covariates

In addition to crude vaccination rates, we estimated vaccination rates adjusted for a range of factors known to be associated with vaccination uptake and occupation [8], and therefore likely to confound the relationship between occupation and vaccination. We adjusted for sex (Male, Female), age (five-year age band), region, ethnicity (White British, Bangladeshi, Black African, Black Caribbean, Chinese, Indian, Mixed, Other, Pakistani, White other), disability status (non-disabled, disabled and limited a little, disabled and limited a lot), highest level of qualification (Level 4+, Level 3, Apprenticeship, Level 2, Level 1, other, no qualification) and pre-existing conditions (1+) based on the QCovid risk model (See Table S2 for more detail).

### Occupation characteristics

We also used occupation-level (Unit group) data on ability to work from home. These data were derived by the Office for National Statistics based on data from the Occupational Information Network (O*NET), which contains information about the features and the nature of the work of the US [9]. Occupations were assigned a score of between one and five to reflect the frequency and importance of different tasks and characteristics to various jobs. Factors considered included whether the job has to be carried out in a specific location; the amount of face-to-face interaction with others; exposure to infections and other hazards; whether the job requires physical activity; and use of tools or protective equipment. An overall score was derived by summing the category scores, which were first rescaled to between zero and one. The final score was also rescaled to between zero and one, with one indicating a high ability to work from home.

### Statistical analyses

First, we estimated the rate of people aged 40 to 64 who had received two doses of a COVID-19 vaccine by occupation. Second, we estimated age standardised vaccination rates for the different unit groups, whereby the age distribution within each group was standardized to the 2013 European Standardised Population.

Third, we examined whether differences were driven by other socio-demographic and clinical factors likely to be associated with vaccination uptake and occupation. We selected factors known to be associated with vaccination uptake in England [8] and to be linked to occupation. We used logistic regression to estimate the odds of not being fully vaccinated (receiving two doses of the vaccine) by occupation, adjusting for sex, age, geographical and sociodemographic characteristics, disability status and pre-existing conditions. We estimated separate models for each occupation so that the odds ratios could be interpreted as the difference in the odds of not being vaccinated for people working in this occupation compared to all other occupations. We estimated unadjusted odds ratios, odds ratios adjusted for age and sex, and fully adjusted odds ratios.

Finally, we investigated the association between vaccination rate and the ability to work from home. We visualised the relationship using a scatter plot and estimated the strength of the association using univariate linear regression models. We also used logistic regression models to estimate the association between working from home and the odds of not being fully vaccinated, after adjusting for other characteristics. We used the same approach as for occupation and estimated standard error clustered at occupation level. Because the working from home score was standardised with a mean of 0 and a standard deviation of 1, the odds ratio for the score can interpreted as the effect of an increase by one standard deviation. Analyses were conducted using R 3.5.

## Results

### Characteristics of study population

Our study population included 14,298,147 adults aged 40 to 64 years who lived in England at the beginning of the pandemic and reported being employed in 2011. 88.2% of people had received two doses of a vaccine against COVID-19 by 31 August 2021; 2.7% had only received one dose, whilst 9.1% had not received any. The average age was 52.8 years and 51.4% were female; 81.5% identified themselves as White British. 21.5% of people had at least one of the health conditions included in the QCovid risk model. Table 1 provides detailed characteristics of the sample.

**Table 1.**
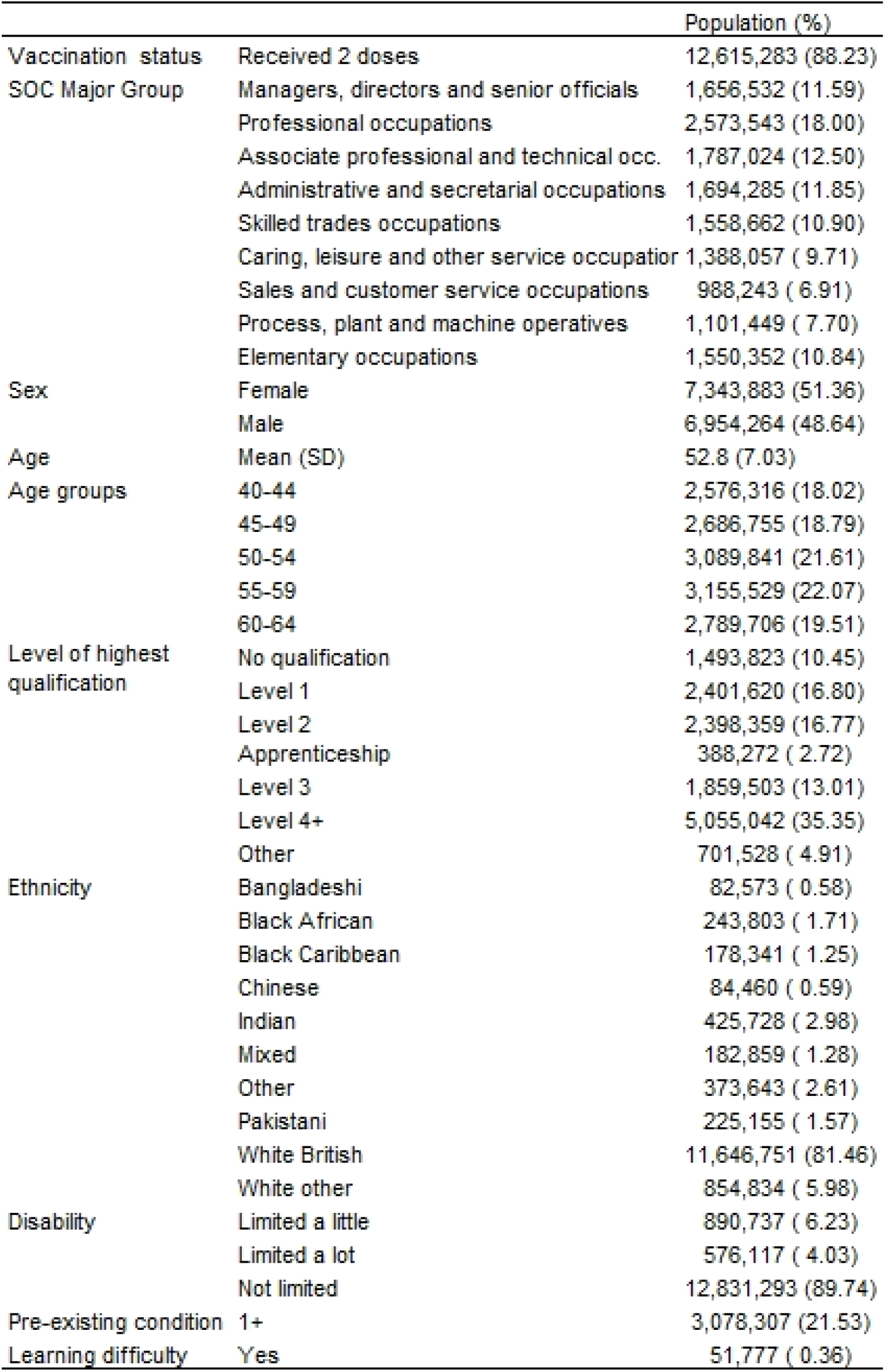
Characteristics of the study population.

|  |  | Population (%) |
| --- | --- | --- |
| Vaccination status | Received 2 doses | 12,615,283 (88.23) |
| SOC Major Group | Managers, directors and senior officials | 1,656,532 (11.59) |
|  | Professional occupations | 2,573,543 (18.00) |
|  | Associate professional and technical occ. | 1,787,024 (12.50) |
|  | Administrative and secretarial occupations | 1,694,285 (11.85) |
|  | Skilled trades occupations | 1,558,662 (10.90) |
|  | Caring, leisure and other service occupation | 1,388,057 (9.71) |
|  | Sales and customer service occupations | 988,243 (6.91) |
|  | Process, plant and machine operatives | 1,101,449 (7.70) |
|  | Elementary occupations | 1,550,352 (10.84) |
| Sex | Female | 7,343,883 (51.36) |
|  | Male | 6,954,264 (48.64) |
| Age | Mean (SD) | 52.8 (7.03) |
| Age groups | 40-44 | 2,576,316 (18.02) |
|  | 45-49 | 2,686,755 (18.79) |
|  | 50-54 | 3,089,841 (21.61) |
|  | 55-59 | 3,155,529 (22.07) |
|  | 60-64 | 2,789,706 (19.51) |
| Level of highest qualification | No qualification | 1,493,823 (10.45) |
|  | Level 1 | 2,401,620 (16.80) |
|  | Level 2 | 2,398,359 (16.77) |
|  | Apprenticeship | 388,272 (2.72) |
|  | Level 3 | 1,859,503 (13.01) |
|  | Level 4+ | 5,055,042 (35.35) |
|  | Other | 701,528 (4.91) |
| Ethnicity | Bangladeshi | 82,573 (0.58) |
|  | Black African | 243,803 (1.71) |
|  | Black Caribbean | 178,341 (1.25) |
|  | Chinese | 84,460 (0.59) |
|  | Indian | 425,728 (2.98) |
|  | Mixed | 182,859 (1.28) |
|  | Other | 373,643 (2.61) |
|  | Pakistani | 225,155 (1.57) |
|  | White British | 11,646,751 (81.46) |
|  | White other | 854,834 (5.98) |
| Disability | Limited a little | 890,737 (6.23) |
|  | Limited a lot | 576,117 (4.03) |
|  | Not limited | 12,831,293 (89.74) |
| Pre-existing condition | 1+ | 3,078,307 (21.53) |
| Learning difficulty | Yes | 51,777 (0.36) |

### Vaccination rates by occupation

As of 31 August 2021, vaccination rates in workers aged 40 to 64 differed by SOC Major Group. Vaccination rates were over 90% in administrative and secretarial occupations (90.8%); professional occupations (90.7%); and managers, directors and senior officials (90.6%). Those working in elementary occupations had the lowest rate of vaccination with 83.1% of people having received two doses of the vaccine (Table 2).

**Table 2.** Vaccination rates by SOC Major Group.

| Occupation (SOC Major group) | Population | No vaccine | First dose only | Two doses |
| --- | --- | --- | --- | --- |
| Managers, directors and senior officials | 1,656,532 | 7.5 (7.4 - 7.5) | 2.0 (2.0 - 2.0) | 90.6 (90.5 - 90.6) |
| Professional occupations | 2,573,543 | 7.6 (7.5 - 7.6) | 1.8 (1.7 - 1.8) | 90.7 (90.6 - 90.7) |
| Associate professional and technical occupations | 1,787,024 | 8.6 (8.6 - 8.7) | 2.1 (2.1 - 2.1) | 89.3 (89.2 - 89.3) |
| Administrative and secretarial occupations | 1,694,285 | 7.1 (7.1 - 7.1) | 2.1 (2.1 - 2.1) | 90.8 (90.8 - 90.9) |
| Skilled trades occupations | 1,558,662 | 10.9 (10.9 - 11.0) | 3.3 (3.2 - 3.3) | 85.8 (85.7 - 85.8) |
| Caring, leisure and other service occupations | 1,388,057 | 9.0 (9.0 - 9.1) | 2.9 (2.9 - 2.9) | 88.1 (88.0 - 88.1) |
| Sales and customer service occupations | 988,243 | 9.6 (9.6 - 9.7) | 3.3 (3.3 - 3.4) | 87.0 (87.0 - 87.1) |
| Process, plant and machine operatives | 1,101,449 | 11.4 (11.3 - 11.4) | 3.3 (3.3 - 3.4) | 85.3 (85.2 - 85.4) |
| Elementary occupations | 1,550,352 | 13.0 (12.9 - 13.1) | 3.9 (3.9 - 4.0) | 83.1 (83.0 - 83.1) |

Note: ONS Public Health Data Asset; Occupation as recorded in 2011 Census

Vaccination rates also differed markedly by occupation unit groups (Table 3 for selected occupation, Supplementary Table S3 for all occupations). Vaccination rates were highest in Senior police officers (95.5% [94.8 – 96.3]), school secretaries (94.8% [94.6 -95.1]), Senior officers in fire, ambulance and prison and related services (94.6 % [94.1 – 95.1]) and police officers (94.4% [94.3 – 94.5%]). The vaccination rate was over 92% in 50 out of 370 occupation unit groups, accounting 1,765,089 (12.3%) workers. High rates of vaccination were found in education workers (particularly primary and nursery education teaching professionals, teaching assistants, senior professional of educational establishments), national and local government administrative occupation, and solicitors.

**Table 3.** Vaccination rates and odds ratios for not being fully vaccinated for SOC unit groups, top and bottom 20.

| Occupation | Population | Number vaccinated | Vaccination rate | Age standardised rate | Unadjusted OR | Fully adjusted OR |
| --- | --- | --- | --- | --- | --- | --- |
| Packers, bottlers, canners and fillers | 72,464 | 55,838 | 77.1 (76.8 - 77.4) | 76.5 (75.9 - 77.1) | 2.24 (2.21 - 2.28) | 1.43 (1.40 - 1.45) |
| Health associate professionals n.e.c. | 21,136 | 16,318 | 77.2 (76.6 - 77.8) | 76.7 (75.5 - 77.9) | 2.22 (2.15 - 2.29) | 3.00 (2.90 - 3.10) |
| Fitness instructors | 18,657 | 14,511 | 77.8 (77.2 - 78.4) | 78.6 (77.3 - 79.9) | 2.14 (2.07 - 2.22) | 2.29 (2.21 - 2.38) |
| Vehicle valeters and cleaners | 14,511 | 11,402 | 78.6 (77.9 - 79.2) | 79.2 (77.8 - 80.7) | 2.05 (1.97 - 2.13) | 1.28 (1.23 - 1.33) |
| Elementary construction occupations | 80,244 | 63,166 | 78.7 (78.4 - 79.0) | 77.7 (77.1 - 78.4) | 2.04 (2.00 - 2.07) | 1.52 (1.49 - 1.55) |
| Waiters and waitresses | 52,218 | 41,224 | 78.9 (78.6 - 79.3) | 81.0 (80.2 - 81.9) | 2.01 (1.96 - 2.05) | 1.18 (1.15 - 1.20) |
| Market and street traders and assistants | 8,831 | 7,084 | 80.2 (79.4 - 81.0) | 77.5 (75.6 - 79.4) | 1.85 (1.76 - 1.95) | 1.45 (1.37 - 1.53) |
| Plasterers | 26,578 | 21,310 | 80.2 (79.7 - 80.7) | 79.4 (78.3 - 80.5) | 1.86 (1.80 - 1.91) | 1.62 (1.57 - 1.67) |
| Scaffolders, staggers and riggers | 14,606 | 11,737 | 80.4 (79.7 - 81.0) | 80.8 (79.4 - 82.3) | 1.83 (1.76 - 1.91) | 1.61 (1.54 - 1.68) |
| Actors, entertainers and presenters | 19,150 | 15,422 | 80.5 (80.0 - 81.1) | 81.0 (79.7 - 82.3) | 1.81 (1.75 - 1.88) | 1.61 (1.55 - 1.68) |
| Industrial cleaning process occupations | 10,974 | 8,862 | 80.8 (80.0 - 81.5) | 78.6 (76.9 - 80.3) | 1.79 (1.70 - 1.87) | 1.40 (1.33 - 1.47) |
| Bar staff | 55,784 | 45,076 | 80.8 (80.5 - 81.1) | 82.5 (81.7 - 83.3) | 1.79 (1.75 - 1.82) | 1.51 (1.47 - 1.54) |
| Leisure and theme park attendants | 4,797 | 3,881 | 80.9 (79.8 - 82.0) | 81.9 (79.3 - 84.5) | 1.77 (1.65 - 1.90) | 1.39 (1.28 - 1.49) |
| Tailors and dressmakers | 8,687 | 7,030 | 80.9 (80.1 - 81.8) | 79.7 (77.8 - 81.6) | 1.77 (1.68 - 1.86) | 1.16 (1.09 - 1.22) |
| Elementary security occupations n.e.c. | 9,977 | 8,099 | 81.2 (80.4 - 81.9) | 80.6 (78.9 - 82.4) | 1.74 (1.65 - 1.83) | 1.40 (1.32 - 1.47) |
| Roofers, roof tilers and slaters | 25,969 | 21,078 | 81.2 (80.7 - 81.6) | 80.3 (79.2 - 81.4) | 1.74 (1.69 - 1.80) | 1.55 (1.50 - 1.60) |
| Fork-lift truck drivers | 39,486 | 32,148 | 81.4 (81.0 - 81.8) | 80.5 (79.7 - 81.4) | 1.71 (1.67 - 1.76) | 1.25 (1.22 - 1.29) |
| Window cleaners | 16,746 | 13,648 | 81.5 (80.9 - 82.1) | 80.0 (78.7 - 81.4) | 1.70 (1.64 - 1.77) | 1.53 (1.47 - 1.60) |
| Food, drink and tobacco process operatives | 90,746 | 73,959 | 81.5 (81.2 - 81.8) | 80.8 (80.3 - 81.4) | 1.71 (1.68 - 1.74) | 1.17 (1.15 - 1.19) |
| Therapy professionals n.e.c. | 11,688 | 9,566 | 81.8 (81.1 - 82.5) | 80.7 (79.0 - 82.4) | 1.66 (1.59 - 1.74) | 2.36 (2.24 - 2.48) |
| .... | .... | .... | .... | .... | .... | .... |
| Human resource managers and directors | 56,846 | 52,844 | 93.0 (92.7 - 93.2) | 92.9 (92.1 - 93.7) | 0.57 (0.55 - 0.59) | 0.72 (0.70 - 0.74) |
| Solicitors | 59,739 | 55,615 | 93.1 (92.9 - 93.3) | 93.5 (92.7 - 94.3) | 0.55 (0.54 - 0.57) | 0.60 (0.58 - 0.62) |
| Chartered surveyors | 33,252 | 30,967 | 93.1 (92.9 - 93.4) | 92.8 (91.7 - 93.8) | 0.55 (0.53 - 0.58) | 0.72 (0.68 - 0.75) |
| Medical radiographers | 11,307 | 10,543 | 93.2 (92.8 - 93.7) | 93.1 (91.4 - 94.9) | 0.54 (0.50 - 0.58) | 0.69 (0.64 - 0.74) |
| Purchasing managers and directors | 24,815 | 23,127 | 93.2 (92.9 - 93.5) | 92.9 (91.7 - 94.1) | 0.55 (0.52 - 0.57) | 0.68 (0.65 - 0.71) |
| Librarians | 9,870 | 9,215 | 93.4 (92.9 - 93.9) | 92.6 (90.6 - 94.6) | 0.53 (0.49 - 0.58) | 0.74 (0.68 - 0.80) |
| Medical secretaries | 33,188 | 31,002 | 93.4 (93.1 - 93.7) | 92.4 (91.3 - 93.6) | 0.53 (0.51 - 0.55) | 0.74 (0.71 - 0.77) |
| Insurance underwriters | 14,945 | 13,966 | 93.4 (93.1 - 93.8) | 93.8 (92.3 - 95.4) | 0.53 (0.49 - 0.56) | 0.60 (0.56 - 0.64) |
| Paramedics | 9,873 | 9,242 | 93.6 (93.1 - 94.1) | 93.7 (91.8 - 95.6) | 0.51 (0.47 - 0.55) | 0.73 (0.67 - 0.79) |
| Health services and public health managers and directors | 29,393 | 27,498 | 93.6 (93.3 - 93.8) | 92.6 (91.4 - 93.8) | 0.52 (0.49 - 0.54) | 0.73 (0.70 - 0.77) |
| Senior professionals of educational establishments | 65,028 | 60,875 | 93.6 (93.4 - 93.8) | 92.8 (92.0 - 93.6) | 0.51 (0.49 - 0.53) | 0.76 (0.73 - 0.78) |
| Taxation experts | 14,574 | 13,652 | 93.7 (93.3 - 94.1) | 93.6 (92.0 - 95.2) | 0.51 (0.47 - 0.54) | 0.62 (0.58 - 0.66) |
| Health care practice managers | 10,620 | 9,960 | 93.8 (93.3 - 94.2) | 92.7 (90.8 - 94.7) | 0.50 (0.46 - 0.54) | 0.71 (0.65 - 0.77) |
| Occupational therapists | 14,845 | 13,918 | 93.8 (93.4 - 94.1) | 93.8 (92.3 - 95.4) | 0.50 (0.47 - 0.53) | 0.69 (0.65 - 0.74) |
| Town planning officers | 5,313 | 4,989 | 93.9 (93.3 - 94.5) | 94.3 (91.6 - 97.0) | 0.49 (0.43 - 0.54) | 0.57 (0.50 - 0.63) |
| Speech and language therapists | 6,360 | 6,003 | 94.4 (93.8 - 95.0) | 94.5 (92.1 - 97.0) | 0.45 (0.40 - 0.50) | 0.61 (0.54 - 0.68) |
| Police officers (sergeant and below) | 108,003 | 101,946 | 94.4 (94.3 - 94.5) | 94.4 (93.8 - 95.0) | 0.44 (0.43 - 0.46) | 0.53 (0.51 - 0.54) |
| Senior officers in fire, ambulance, prison and related services | 8,157 | 7,714 | 94.6 (94.1 - 95.1) | 93.4 (91.0 - 95.9) | 0.43 (0.39 - 0.47) | 0.64 (0.58 - 0.71) |
| School secretaries | 34,524 | 32,739 | 94.8 (94.6 - 95.1) | 92.8 (91.6 - 94.0) | 0.41 (0.39 - 0.43) | 0.62 (0.59 - 0.65) |
| Senior police officers | 2,775 | 2,651 | 95.5 (94.8 - 96.3) | 94.2 (88.6 - 99.8) | 0.35 (0.29 - 0.42) | 0.60 (0.50 - 0.72) |

Note: ONS Public Health Data Asset; Results for all SOC unit groups are presented in Supplementary Table S3. Vaccination rate is the rate of people fully vaccinated. The outcome of the logistic regression models is not being fully vaccinated as of 31 August 2021. The reference category for the ORs is all other occupations. Fully adjusted models include sex, five-year age bands, ethnicity, region, education, disability, pre-existing condition.

Vaccination rates were lower than 80% in just six occupations, accounting for 259,230 (1.8%) people in our dataset. These included packers, bottlers, canners and fillers (77.1% [76.8 – 77.4]), health associated professionals such as acupuncturists, Chiropractors and osteopaths (77.2% [76.6 – 77.8]), fitness instructors (77.8% [77.2 – 78.4]), vehicle valeters and cleaners (78.6% [77.9 – 79.2]), elementary construction occupations (78.7% [78.4 – 79.0]), and waiters and waitresses (78.9% [78.6 – 79.3]).

The vaccination rates were lower than 85% in 70 out of 370 occupations. 2.705,811 (18.9%) people worked in these occupations. The larger occupations (eg with more than 100,000 workers) with low vaccination rates included chefs (82.6% [82.3 – 82.8]), cleaners and domestics (82.7% [82.6 – 82.9]), people working in elementary storage occupations (82.8% [82.6 – 83.0]), taxi and cab drivers (83.3% [83.1 – 83.6]), van drivers (84.3% [84.1 – 84.5]), and carpenters and joiners (84.3% [84.1 – 84.6]). Low rates of vaccination were also found in kitchen and catering assistants (85.2% [85.0 – 85.4], care workers and home carers (85.8% [85.6 – 85.9]) and sales and retail assistant (86.4% [86.3 – 86.5]), three large occupational groups (over 1 million of workers in our sample) which involve working with the public.

Amongst health care workers, there was some variation in vaccination rates. Most unit groups amongst health professionals had vaccination rates above average, with 91.7% of nurses and midwives being fully vaccinated and 89.4% of medical practioners. Vaccine coverage was low in health associated professionals not elsewhere classified group (e.g. acupuncturists, homeopaths and hypnotherapists) and therapy professionals not elsewhere classified (e.g. art therapists, chiropractor, Cognitive behavioural therapist and dance movement therapists). Vaccination rates were also low in care workers and home carers

Standardising for age or adjusting further for other factors likely to be linked to occupation and vaccination did not substantially alter the results (See Supplementary Table S3). Out of the 140 occupations which had an unadjusted OR for not being fully vaccinated significantly greater than one, only 7 changed to being significantly lower than one after adjusting for socio-demographic characteristics and pre-existing conditions. These included restaurant and catering managers, chefs, and taxi drivers. For another 23 occupations, the adjusted OR was not different from one. Adjusted OR for not being fully vaccinated were highest in health associated professionals not elsewhere classified (3.00 [2.90 – 3.10]), therapy professionals not elsewhere classified (2.37 [2.24 – 2.48]) and fitness instructors (2.29 [2.21 – 2.38]).

Out of the 189 occupations which had an unadjusted OR significantly lower than one, only 7 changed to being significantly greater than one after adjustment, including receptionists and metal workers. A further 27 became not different from one after adjustment. The lowest odds ratios for not being fully vaccinated were found in police officers (0.53 [0.51 – 0.54]), tow planning officers (0.57 [0.50 -0.63]), and solicitors (0.60 [0.58 – 0.62]).

Results were similar when looking at people who had not received any vaccine (See Supplementary Table S4), suggesting that the results are not driven by people working in some occupation being more likely not have been eligible to receive their second dose.

### Vaccination rates and job characteristics

Vaccination rates were associated with the ability to work from home, as assessed based on O*NET data. Figure 1 shows that vaccination rates tend to be higher in occupations in which working from home is more easily possible. The association was strong, with a R-squared of 0.208 and a F-statistics of 95.4. Results from a logistic regression model suggests that one standard deviation increase in the working from home score is associated with 0.46 [0.37 - 0.58] times lower odds of not being fully vaccinated. Adjusting for geographical factors, socio-demographic characteristics and health reduced the ORs did not substantially affect the results (adjusted ORs 0.59 [0.48 -0.69]), indicating that working from home is independently associated with a reduction in the odds of not being vaccinated.

**Figure 1.**
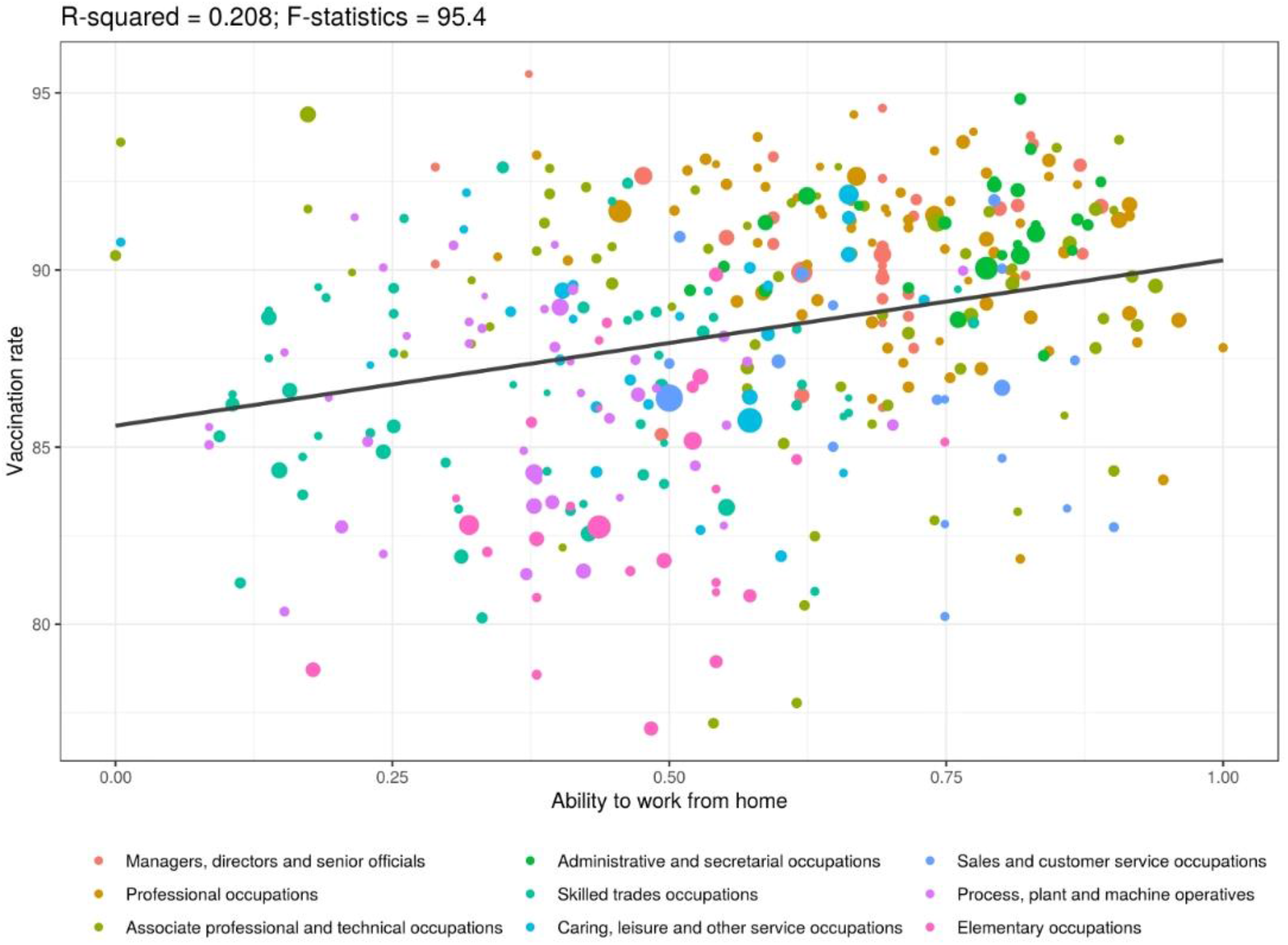
Association between vaccination rates and ability to work from home at unit group level.

## Discussion

### Main findings of this study

Using whole population level linked data in England, our analysis shows that the vaccination rates of adults aged 40 to 64 years and over differed markedly by occupation. Vaccination rates were high in administrative and secretarial occupations, professional occupations and managers, directors and senior officials and low in people working in elementary occupations. Vaccination rates were also associated with the ability to work from home, with vaccine uptake being higher in occupations which can be performed from home (this relationship remained when sociodemographic characteristics were adjusted for).

### Comparison with other studies

Few studies have investigated how COVID-19 vaccination coverage varies by occupation. Several studies investigated vaccine hesitancy and vaccine coverage in health care workers, highlighting some differences between different job roles, with doctors having slightly higher vaccination coverage and lower hesitancy than midwives and nurses [10, 11]. We found no substantial differences in vaccination rates between nurses and medical practitioners, but found much lower vaccination rates in health associated professionals such as acupuncturists, Chiropractors and osteopaths, and in care workers and home carers. The difference between our results and those from studies focused on health care workers may stem from the age range selected (40 to 64) and recent migrants being excluded from our dataset. Migrants make up about a fifth of the health and social care workforce [12] and people born outside of the UK tend to have lower vaccination coverage [13].

Our findings that vaccination rates are higher among managers, directors and senior officials and in people working in professional occupations, compared with those working in elementary occupations, are in line with studies showing that vaccination rates in the UK and the US are higher in more wealthy areas and amongst people from higher socio-economic status [13, 3, 14, 4]. Our results are also consistent with a study from the US showing that vaccine hesitancy varied widely by occupation category, with hesitancy being low in people working in professional occupations, especially in the life, physical and social sciences and particularly high in people working in construction and extraction [15].

### Strengths and limitations

A major strength of this study is the use of nationwide linked popula1tion-level data from clinical records and the 2011 Census. This study is the first to examine how vaccination rates vary by occupation using population-level data. Because information on occupation is not collected in electronic health records, we used data from the Census to assess people’s occupation. Some surveys collect data on both occupations and vaccination but face the issue that non-response is likely correlated with the propensity to be vaccinated. Having population level data based on electronic health records and the Census, which is mandatory and has a high response rate, we were able to accurately estimate vaccination rates for detailed occupational groups.

The main limitation of our study is that the information on occupation is nine years out of date. Our exposure is therefore likely to be misclassified for a proportion of people, because they have left the labour force or changed occupation since 2011, especially during the pandemic. To mitigate measurement error, we restricted our analysis to people aged 40-64 years, who have a relatively high occupational stability, as shown in an analysis of a large longitudinal household survey [7]. Exposure misclassification is nonetheless likely result in underestimating the differences in vaccination rates between occupations, especially if the exposure misclassification is random. Turnout rate may be greater in some occupations than others, especially in elementary occupations where vaccination rates are lower, which could lead to underestimate the differences in vaccination rates. However, over two-third of people aged 40 to 64 in 2019 were in the same major group as in 2011 [7]. The bias is likely to be smaller in the estimated vaccination rates by major group than by unit group.

An additional limitation is that, because the Public Health Data Asset was based on the 2011 Census, it excluded people who were living in England in 2011 but who did not take part in the Census, as well as respondents who could not be linked to the 2011-2013 NHS patient register and recent migrants. As a result, we excluded 5.4% of vaccinated people who could not be linked to the PHDA.

### Mechanisms

Our analysis highlights substantial differences in vaccination coverage between occupations, with vaccination rates being higher in professional occupations and managers, directors and senior officials and lower in people working in elementary occupations. These differences are only partially explained by other factors, including education, suggesting that some occupations are independently associated with vaccination status. People working in elementary occupations may have little job control and therefore may find it more difficult to attend vaccination or may be put off by the potential side effects, which could prevent them from working. Existing evidence suggests that occupational characteristics, such as job strain, can affect health behaviours, such as compliance with treatment [16]. Vaccine hesitancy may also be driving differences in uptake. The drivers of vaccine hesitancy are complex, and occupation may play a part. Indeed, we find that vaccine coverage is low in health associated professionals such as acupuncturists and osteopaths, and therapy professionals such as art therapist, chiropractor, cognitive behavioural therapists and dance movement therapists. They may be more likely to reject mainstream medicine than other groups. The rates were also low in fitness instructors, who may be more likely to believe they are very healthy and therefore may not need the vaccine.

## Conclusion

In our study population of older worker adults in England, 88.2% had received two doses of a vaccine against COVID-19 by 31 August 2021 – a public health triumph. However, many occupations with lower vaccination rates involved contact with the public (taxi and cab drivers, sales and retail assistant, waiters and waitresses) or with vulnerable people (care workers and home carers). Therefore, increasing vaccination coverage in these occupations would be crucial to help protect the public and control infection. Our results also show that vaccination rates are associated with the ability to work from home, with vaccination rates typically being higher in occupations which can be done from home. Therefore, policies such as ‘work from home if you can’ may only have limited impact on hospitalisations and deaths, as the vaccination rates are higher in the occupations that can be done from home and lower in those which cannot. Efforts should be made to increase vaccination rates in occupations that cannot be done from home. This includes many elementary occupations where it may be difficult to get time off work to be vaccinated, or where workers may not have the spare time or resources to access vaccination or are worried about missing work because of the side effects of the vaccines. As these are some of the more deprived groups in society, increasing vaccination rates in these groups would help reduce inequalities in vaccination coverage. Our results could also help inform future vaccination strategy and future models of COVID-19 transmission.

## Data Availability

The ONS Public Health Linked Data Asset will be made available on the ONS Secure Research Service for Accredited researchers. Researchers can apply for accreditation through the Research Accreditation Service.

## Ethical Approval Statement

Ethical approval was obtained from the National Statistician’s Data Ethics Advisory Committee (NSDEC(20)12).

## Authorship statement

Study conceptualisation was led by VN and TD. VN, TD and DA contributed to the development of the research question, study design, with development of statistical aspects led by TD and VN. TD and VN were involved in data specification, curation and collection. TD and VN conducted and checked the statistical analyses. VN, TD, KF, RE, MG, NP, MvT, contributed to the interpretation of the results. VN wrote the first draft of the paper. TD, MG, KF, RE, NP, MvT contributed to the critical revision of the manuscript for important intellectual content and approved the final version of the manuscript. VN had full access to all data in the study and takes responsibility of the integrity of the data and the accuracy of the data analysis. VN affirms that the manuscript is an honest, accurate, and transparent account of the study being reported; that no important aspects of the study have been omitted; and that any discrepancies from the study as planned have been explained

## Supplementary material

**Table S1.**
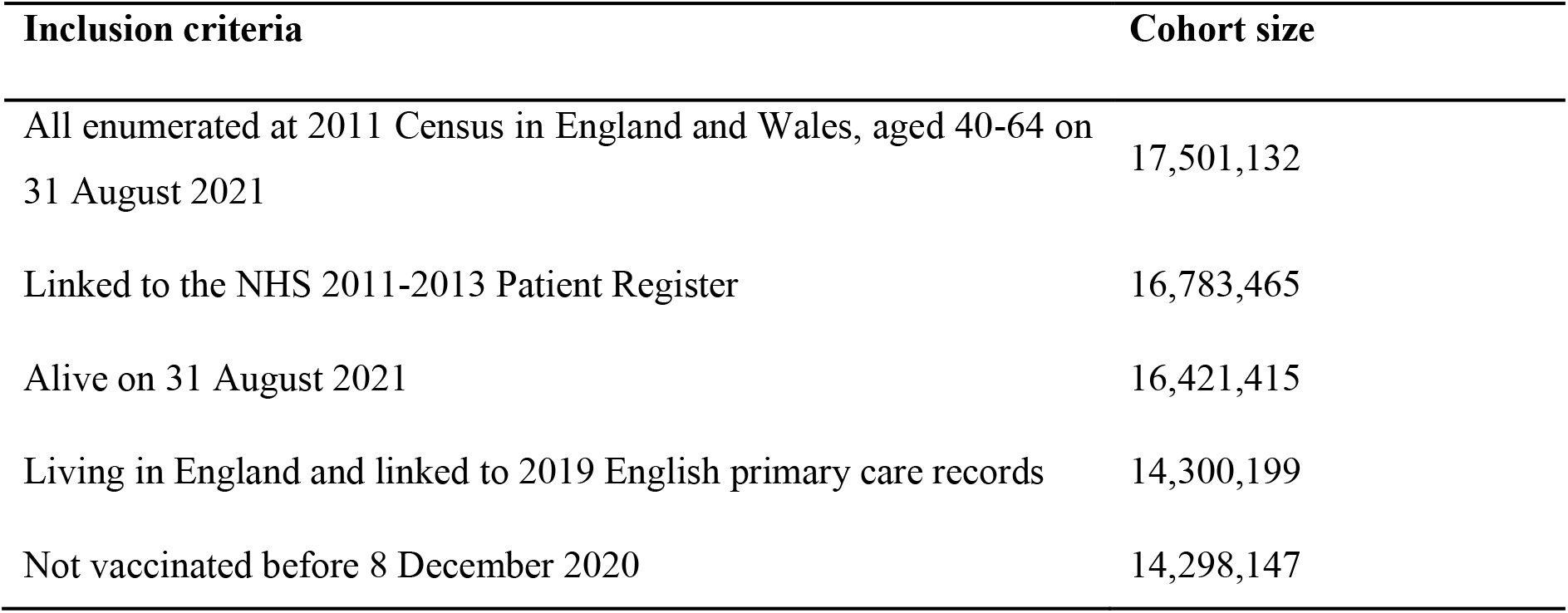
Sample selection and number of participants.

| Inclusion criteria | Cohort size |
| --- | --- |
| All enumerated at 2011 Census in England and Wales, aged 40-64 on 31 August 2021 | 17,501,132 |
| Linked to the NHS 2011-2013 Patient Register | 16,783,465 |
| Alive on 31 August 2021 | 16,421,415 |
| Living in England and linked to 2019 English primary care records | 14,300,199 |
| Not vaccinated before 8 December 2020 | 14,298,147 |

**Table S2.**
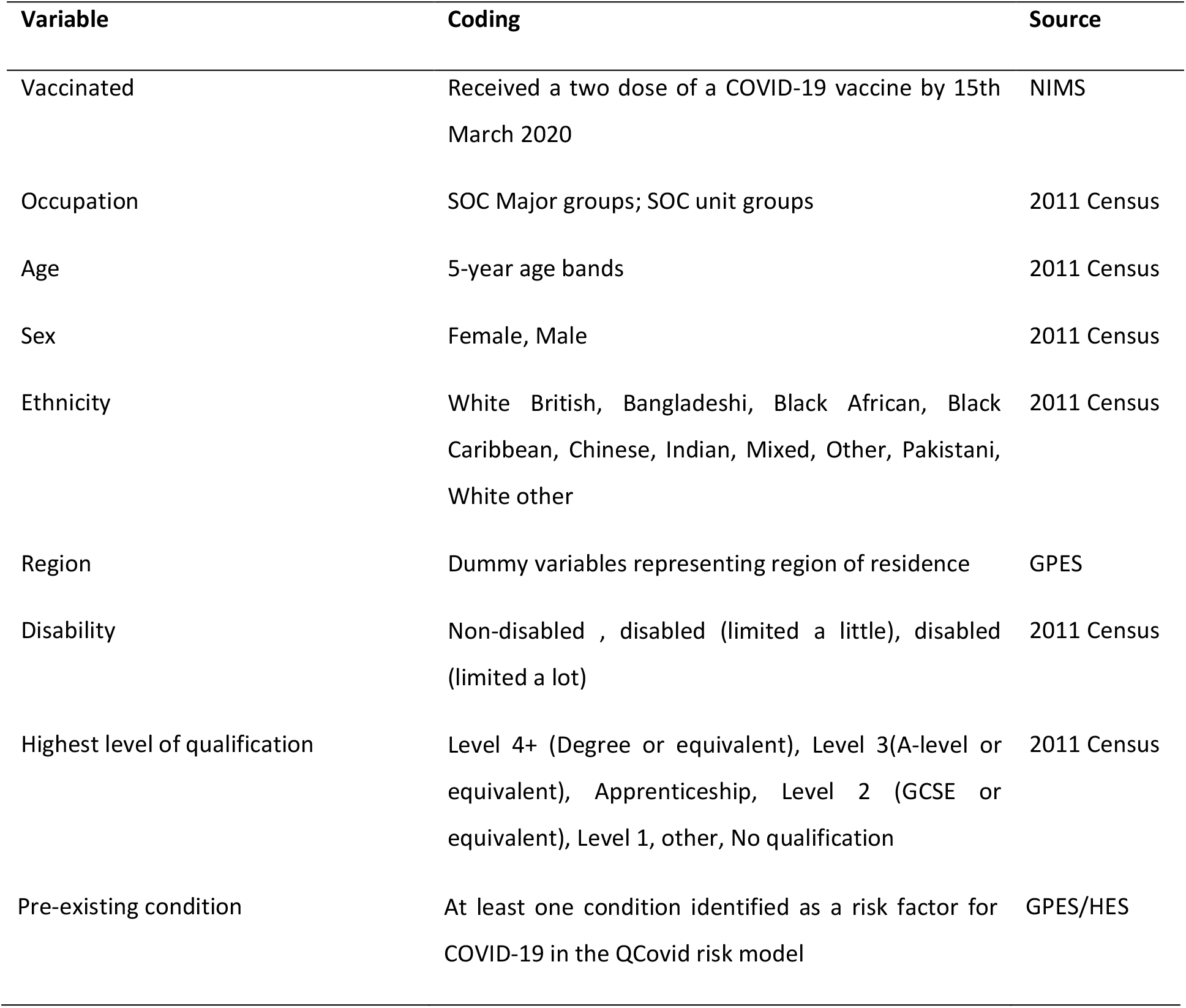
Variables used in the analyses.

**Table S3.** Vaccination rates and odds ratios for SOC unit groups.

| Occupation | Population | Number vaccinated | Vaccination rate | Age standardised rate | Unadjusted OR | Age-sex adjusted OR | Fully adjusted OR |
| --- | --- | --- | --- | --- | --- | --- | --- |
| Chief executives and senior officials | 23,367 | 20,514 | 87.8 (87.4 - 88.2) | 85.3 (84.0 - 86.6) | 1.04 (1.00 - 1.08) | 1.16 (1.12 - 1.21) | 1.31 (1.26 - 1.36) |
| Elected officers and representatives | 2,640 | 2,400 | 90.9 (89.8 - 92.0) | 89.5 (85.6 - 93.5) | 0.75 (0.66 - 0.86) | 0.85 (0.75 - 0.97) | 0.87 (0.76 - 0.99) |
| Production managers and directors in manufacturing | 157,327 | 145,776 | 92.7 (92.5 - 92.8) | 91.6 (91.1 - 92.1) | 0.59 (0.58 - 0.60) | 0.60 (0.59 - 0.61) | 0.76 (0.75 - 0.78) |
| Production managers and directors in construction | 98,624 | 89,664 | 90.9 (90.7 - 91.1) | 89.3 (88.6 - 89.9) | 0.75 (0.73 - 0.76) | 0.75 (0.74 - 0.77) | 0.89 (0.87 - 0.91) |
| Production managers and directors in mining and energy | 9,115 | 8,439 | 92.6 (92.0 - 93.1) | 92.1 (90.1 - 94.1) | 0.60 (0.56 - 0.65) | 0.59 (0.55 - 0.64) | 0.81 (0.74 - 0.87) |
| Financial managers and directors | 91,614 | 84,097 | 91.8 (91.6 - 92.0) | 91.4 (90.8 - 92.0) | 0.67 (0.65 - 0.68) | 0.65 (0.64 - 0.67) | 0.76 (0.74 - 0.78) |
| Marketing and sales directors | 60,082 | 55,170 | 91.8 (91.6 - 92.0) | 91.2 (90.4 - 91.9) | 0.67 (0.65 - 0.69) | 0.65 (0.63 - 0.67) | 0.81 (0.79 - 0.84) |
| Purchasing managers and directors | 24,815 | 23,127 | 93.2 (92.9 - 93.5) | 92.9 (91.7 - 94.1) | 0.55 (0.52 - 0.57) | 0.53 (0.50 - 0.56) | 0.68 (0.65 - 0.71) |
| Advertising and public relations directors | 15,633 | 14,045 | 89.8 (89.4 - 90.3) | 90.2 (88.7 - 91.7) | 0.85 (0.80 - 0.89) | 0.75 (0.72 - 0.79) | 0.92 (0.87 - 0.97) |
| Human resource managers and directors | 56,846 | 52,844 | 93.0 (92.7 - 93.2) | 92.9 (92.1 - 93.7) | 0.57 (0.55 - 0.59) | 0.57 (0.55 - 0.59) | 0.72 (0.70 - 0.74) |
| Information technology and telecommunications directors | 36,476 | 32,995 | 90.5 (90.2 - 90.8) | 89.7 (88.7 - 90.7) | 0.79 (0.76 - 0.82) | 0.75 (0.72 - 0.78) | 0.96 (0.93 - 1.00) |
| Functional managers and directors n.e.c. | 41,145 | 37,301 | 90.7 (90.4 - 90.9) | 89.7 (88.7 - 90.6) | 0.77 (0.75 - 0.80) | 0.81 (0.78 - 0.84) | 0.93 (0.90 - 0.96) |
| Financial institution managers and directors | 82,989 | 76,129 | 91.7 (91.5 - 91.9) | 91.4 (90.7 - 92.0) | 0.67 (0.66 - 0.69) | 0.65 (0.63 - 0.66) | 0.72 (0.70 - 0.74) |
| Managers and directors in transport and distribution | 47,884 | 43,803 | 91.5 (91.2 - 91.7) | 90.4 (89.5 - 91.3) | 0.70 (0.68 - 0.72) | 0.70 (0.68 - 0.72) | 0.79 (0.76 - 0.81) |
| Managers and directors in storage and warehousing | 37,533 | 34,056 | 90.7 (90.4 - 91.0) | 90.1 (89.1 - 91.0) | 0.76 (0.74 - 0.79) | 0.72 (0.69 - 0.74) | 0.77 (0.74 - 0.80) |
| Officers in armed forces | 4,829 | 4,293 | 88.9 (88.0 - 89.8) | 87.7 (85.0 - 90.4) | 0.94 (0.86 - 1.02) | 0.92 (0.84 - 1.00) | 1.24 (1.13 - 1.37) |
| Senior police officers | 2,775 | 2,651 | 95.5 (94.8 - 96.3) | 94.2 (88.6 - 99.8) | 0.35 (0.29 - 0.42) | 0.41 (0.34 - 0.49) | 0.60 (0.50 - 0.72) |
| Senior officers in fire, ambulance, prison and related services | 8,157 | 7,714 | 94.6 (94.1 - 95.1) | 93.4 (91.0 - 95.9) | 0.43 (0.39 - 0.47) | 0.48 (0.43 - 0.53) | 0.64 (0.58 - 0.71) |
| Health services and public health managers and directors | 29,393 | 27,498 | 93.6 (93.3 - 93.8) | 92.6 (91.4 - 93.8) | 0.52 (0.49 - 0.54) | 0.60 (0.58 - 0.63) | 0.73 (0.70 - 0.77) |
| Social services managers and directors | 25,084 | 22,957 | 91.5 (91.2 - 91.9) | 90.8 (89.5 - 92.0) | 0.69 (0.66 - 0.73) | 0.81 (0.77 - 0.85) | 0.95 (0.91 - 0.99) |
| Managers and directors in retail and wholesale | 256,937 | 231,084 | 89.9 (89.8 - 90.1) | 89.7 (89.3 - 90.0) | 0.84 (0.83 - 0.85) | 0.80 (0.79 - 0.81) | 0.86 (0.85 - 0.87) |
| Managers and proprietors in agriculture and horticulture | 8,429 | 7,831 | 92.9 (92.4 - 93.5) | 91.5 (89.4 - 93.7) | 0.57 (0.53 - 0.62) | 0.60 (0.55 - 0.65) | 0.77 (0.71 - 0.84) |
| Managers and proprietors in forestry, fishing and related services | 7,189 | 6,482 | 90.2 (89.5 - 90.9) | 89.1 (86.8 - 91.3) | 0.82 (0.76 - 0.88) | 0.87 (0.80 - 0.94) | 1.10 (1.01 - 1.19) |
| Hotel and accommodation managers and proprietors | 25,317 | 22,454 | 88.7 (88.3 - 89.1) | 87.8 (86.6 - 89.0) | 0.96 (0.92 - 0.99) | 1.00 (0.96 - 1.04) | 0.97 (0.93 - 1.01) |
| Restaurant and catering establishment managers and proprietors | 59,946 | 51,167 | 85.4 (85.1 - 85.6) | 85.5 (84.7 - 86.2) | 1.29 (1.26 - 1.32) | 1.20 (1.18 - 1.23) | 0.91 (0.89 - 0.93) |
| Publicans and managers of licensed premises | 32,977 | 29,456 | 89.3 (89.0 - 89.7) | 88.6 (87.5 - 89.6) | 0.90 (0.87 - 0.93) | 0.93 (0.90 - 0.97) | 1.02 (0.98 - 1.05) |
| Leisure and sports managers | 31,507 | 28,100 | 89.2 (88.8 - 89.5) | 89.4 (88.3 - 90.4) | 0.91 (0.88 - 0.94) | 0.82 (0.79 - 0.85) | 0.97 (0.94 - 1.01) |
| Travel agency managers and proprietors | 7,617 | 6,865 | 90.1 (89.5 - 90.8) | 90.2 (88.1 - 92.3) | 0.82 (0.76 - 0.89) | 0.78 (0.72 - 0.84) | 0.85 (0.79 - 0.92) |
| Health care practice managers | 10,620 | 9,960 | 93.8 (93.3 - 94.2) | 92.7 (90.8 - 94.7) | 0.50 (0.46 - 0.54) | 0.61 (0.56 - 0.66) | 0.71 (0.65 - 0.77) |
| Residential, day and domiciliary care managers and proprietors | 29,250 | 26,906 | 92.0 (91.7 - 92.3) | 91.2 (90.1 - 92.4) | 0.65 (0.63 - 0.68) | 0.76 (0.73 - 0.80) | 0.91 (0.87 - 0.95) |
| Property, housing and estate managers | 69,897 | 62,749 | 89.8 (89.5 - 90.0) | 88.7 (88.0 - 89.4) | 0.85 (0.83 - 0.87) | 0.91 (0.89 - 0.93) | 0.99 (0.96 - 1.01) |
| Garage managers and proprietors | 15,726 | 14,141 | 89.9 (89.5 - 90.4) | 88.1 (86.6 - 89.7) | 0.84 (0.80 - 0.88) | 0.84 (0.80 - 0.88) | 0.99 (0.94 - 1.04) |
| Hairdressing and beauty salon managers and proprietors | 7,620 | 6,562 | 86.1 (85.3 - 86.9) | 86.4 (84.2 - 88.5) | 1.21 (1.13 - 1.29) | 1.22 (1.14 - 1.30) | 1.35 (1.27 - 1.45) |
| Shopkeepers and proprietors ? wholesale and retail | 86,453 | 74,743 | 86.5 (86.2 - 86.7) | 84.4 (83.7 - 85.0) | 1.18 (1.15 - 1.20) | 1.29 (1.26 - 1.31) | 1.14 (1.12 - 1.17) |
| Waste disposal and environmental services managers | 7,067 | 6,255 | 88.5 (87.8 - 89.3) | 87.2 (84.9 - 89.4) | 0.97 (0.90 - 1.05) | 0.97 (0.90 - 1.04) | 1.10 (1.02 - 1.18) |
| Managers and proprietors in other services n.e.c. | 143,652 | 129,916 | 90.4 (90.3 - 90.6) | 89.7 (89.2 - 90.2) | 0.79 (0.78 - 0.80) | 0.81 (0.79 - 0.82) | 0.94 (0.93 - 0.96) |
| Chemical scientists | 5,772 | 5,285 | 91.6 (90.8 - 92.3) | 91.7 (89.2 - 94.2) | 0.69 (0.63 - 0.76) | 0.61 (0.56 - 0.68) | 0.79 (0.72 - 0.87) |
| Biological scientists and biochemists | 22,634 | 20,691 | 91.4 (91.1 - 91.8) | 91.5 (90.3 - 92.8) | 0.70 (0.67 - 0.74) | 0.66 (0.63 - 0.70) | 0.78 (0.75 - 0.82) |
| Physical scientists | 4,712 | 4,146 | 88.0 (87.1 - 88.9) | 88.3 (85.6 - 91.0) | 1.02 (0.94 - 1.12) | 0.89 (0.81 - 0.97) | 1.11 (1.02 - 1.22) |
| Social and humanities scientists | 3,611 | 3,239 | 89.7 (88.7 - 90.7) | 89.8 (86.7 - 92.9) | 0.86 (0.77 - 0.96) | 0.79 (0.71 - 0.88) | 0.97 (0.87 - 1.09) |
| Natural and social science professionals n.e.c. | 30,728 | 26,639 | 86.7 (86.3 - 87.1) | 88.1 (87.0 - 89.2) | 1.15 (1.11 - 1.19) | 0.94 (0.90 - 0.97) | 1.00 (0.97 - 1.03) |
| Civil engineers | 27,649 | 24,534 | 88.7 (88.4 - 89.1) | 88.7 (87.6 - 89.8) | 0.95 (0.92 - 0.99) | 0.83 (0.80 - 0.86) | 0.96 (0.92 - 1.00) |
| Mechanical engineers | 40,123 | 35,769 | 89.1 (88.8 - 89.5) | 88.7 (87.8 - 89.6) | 0.91 (0.88 - 0.94) | 0.82 (0.79 - 0.84) | 1.11 (1.07 - 1.14) |
| Electrical engineers | 12,649 | 11,459 | 90.6 (90.1 - 91.1) | 89.7 (88.0 - 91.3) | 0.78 (0.73 - 0.83) | 0.72 (0.68 - 0.77) | 0.98 (0.93 - 1.05) |
| Electronics engineers | 14,470 | 12,992 | 89.8 (89.3 - 90.3) | 88.9 (87.3 - 90.5) | 0.85 (0.81 - 0.90) | 0.81 (0.77 - 0.85) | 1.04 (0.98 - 1.10) |
| Design and development engineers | 24,802 | 22,667 | 91.4 (91.0 - 91.7) | 91.3 (90.1 - 92.5) | 0.71 (0.68 - 0.74) | 0.61 (0.59 - 0.64) | 0.85 (0.81 - 0.89) |
| Production and process engineers | 14,734 | 13,437 | 91.2 (90.7 - 91.7) | 90.8 (89.3 - 92.3) | 0.72 (0.68 - 0.77) | 0.66 (0.62 - 0.70) | 0.83 (0.78 - 0.88) |
| Engineering professionals n.e.c. | 45,537 | 41,191 | 90.5 (90.2 - 90.7) | 90.2 (89.4 - 91.1) | 0.79 (0.77 - 0.82) | 0.71 (0.69 - 0.73) | 0.94 (0.91 - 0.97) |
| IT specialist managers | 96,216 | 88,367 | 91.8 (91.7 - 92.0) | 91.7 (91.1 - 92.3) | 0.66 (0.65 - 0.68) | 0.60 (0.59 - 0.61) | 0.74 (0.72 - 0.75) |
| IT project and programme managers | 26,939 | 24,658 | 91.5 (91.2 - 91.9) | 91.2 (90.1 - 92.4) | 0.69 (0.66 - 0.72) | 0.65 (0.62 - 0.67) | 0.79 (0.76 - 0.83) |
| IT business analysts, architects and systems designers | 48,073 | 43,509 | 90.5 (90.2 - 90.8) | 90.6 (89.7 - 91.4) | 0.79 (0.76 - 0.81) | 0.68 (0.66 - 0.70) | 0.82 (0.79 - 0.84) |
| Programmers and software development professionals | 86,799 | 76,889 | 88.6 (88.4 - 88.8) | 89.3 (88.6 - 89.9) | 0.97 (0.95 - 0.99) | 0.77 (0.76 - 0.79) | 0.90 (0.88 - 0.92) |
| Web design and development professionals | 21,445 | 18,030 | 84.1 (83.6 - 84.6) | 85.4 (83.9 - 86.8) | 1.42 (1.37 - 1.47) | 1.05 (1.01 - 1.09) | 1.20 (1.15 - 1.25) |
| Information technology and telecommunications professionals n.e.c. | 80,010 | 71,030 | 88.8 (88.6 - 89.0) | 89.0 (88.3 - 89.6) | 0.95 (0.93 - 0.97) | 0.81 (0.80 - 0.83) | 0.92 (0.90 - 0.94) |
| Conservation professionals | 4,264 | 3,962 | 92.9 (92.1 - 93.7) | 93.1 (90.2 - 96.0) | 0.57 (0.51 - 0.64) | 0.52 (0.46 - 0.59) | 0.79 (0.70 - 0.89) |
| Environment professionals | 11,245 | 10,312 | 91.7 (91.2 - 92.2) | 92.1 (90.3 - 93.9) | 0.68 (0.63 - 0.73) | 0.59 (0.55 - 0.63) | 0.80 (0.74 - 0.85) |
| Research and development managers | 18,001 | 16,594 | 92.2 (91.8 - 92.6) | 92.3 (90.9 - 93.8) | 0.64 (0.60 - 0.67) | 0.58 (0.55 - 0.61) | 0.73 (0.69 - 0.77) |
| Medical practitioners | 97,801 | 87,393 | 89.4 (89.2 - 89.6) | 89.5 (88.9 - 90.1) | 0.89 (0.87 - 0.91) | 0.82 (0.81 - 0.84) | 0.87 (0.85 - 0.89) |
| Psychologists | 11,497 | 10,499 | 91.3 (90.8 - 91.8) | 91.6 (89.8 - 93.4) | 0.71 (0.67 - 0.76) | 0.66 (0.62 - 0.70) | 0.78 (0.73 - 0.83) |
| Pharmacists | 20,123 | 18,139 | 90.1 (89.7 - 90.6) | 90.6 (89.3 - 91.9) | 0.82 (0.78 - 0.86) | 0.76 (0.72 - 0.79) | 0.77 (0.73 - 0.81) |
| Ophthalmic opticians | 6,369 | 5,862 | 92.0 (91.4 - 92.7) | 92.4 (90.0 - 94.8) | 0.65 (0.59 - 0.71) | 0.59 (0.54 - 0.65) | 0.71 (0.65 - 0.78) |
| Dental practitioners | 17,182 | 15,510 | 90.3 (89.8 - 90.7) | 90.3 (88.9 - 91.7) | 0.81 (0.77 - 0.85) | 0.77 (0.73 - 0.81) | 0.75 (0.71 - 0.79) |
| Veterinarians | 6,876 | 6,214 | 90.4 (89.7 - 91.1) | 90.8 (88.5 - 93.1) | 0.80 (0.74 - 0.87) | 0.71 (0.66 - 0.77) | 0.80 (0.74 - 0.87) |
| Medical radiographers | 11,307 | 10,543 | 93.2 (92.8 - 93.7) | 93.1 (91.4 - 94.9) | 0.54 (0.50 - 0.58) | 0.55 (0.51 - 0.59) | 0.69 (0.64 - 0.74) |
| Podiatrists | 6,281 | 5,834 | 92.9 (92.2 - 93.5) | 92.7 (90.1 - 95.2) | 0.57 (0.52 - 0.63) | 0.68 (0.61 - 0.75) | 1.00 (0.91 - 1.11) |
| Health professionals n.e.c. | 12,302 | 11,166 | 90.8 (90.3 - 91.3) | 90.7 (89.1 - 92.4) | 0.76 (0.72 - 0.81) | 0.78 (0.73 - 0.83) | 0.93 (0.87 - 0.99) |
| Physiotherapists | 18,805 | 17,453 | 92.8 (92.4 - 93.2) | 93.1 (91.7 - 94.5) | 0.58 (0.55 - 0.61) | 0.54 (0.51 - 0.57) | 0.73 (0.69 - 0.77) |
| Occupational therapists | 14,845 | 13,918 | 93.8 (93.4 - 94.1) | 93.8 (92.3 - 95.4) | 0.50 (0.47 - 0.53) | 0.51 (0.48 - 0.54) | 0.69 (0.65 - 0.74) |
| Speech and language therapists | 6,360 | 6,003 | 94.4 (93.8 - 95.0) | 94.5 (92.1 - 97.0) | 0.45 (0.40 - 0.50) | 0.45 (0.40 - 0.50) | 0.61 (0.54 - 0.68) |
| Therapy professionals n.e.c. | 11,688 | 9,566 | 81.8 (81.1 - 82.5) | 80.7 (79.0 - 82.4) | 1.66 (1.59 - 1.74) | 1.91 (1.82 - 2.00) | 2.36 (2.24 - 2.48) |
| Nurses | 317,899 | 291,384 | 91.7 (91.6 - 91.8) | 91.3 (90.9 - 91.6) | 0.68 (0.67 - 0.69) | 0.77 (0.76 - 0.78) | 0.86 (0.85 - 0.87) |
| Midwives | 19,622 | 17,989 | 91.7 (91.3 - 92.1) | 91.1 (89.7 - 92.5) | 0.68 (0.65 - 0.72) | 0.81 (0.77 - 0.86) | 0.98 (0.93 - 1.03) |
| Higher education teaching professionals | 62,012 | 54,982 | 88.7 (88.4 - 88.9) | 87.8 (87.0 - 88.5) | 0.96 (0.93 - 0.98) | 1.01 (0.99 - 1.04) | 1.06 (1.03 - 1.09) |
| Further education teaching professionals | 45,292 | 40,361 | 89.1 (88.8 - 89.4) | 88.4 (87.5 - 89.3) | 0.92 (0.89 - 0.94) | 1.00 (0.97 - 1.03) | 1.23 (1.20 - 1.27) |
| Secondary education teaching professionals | 184,359 | 168,764 | 91.5 (91.4 - 91.7) | 91.7 (91.3 - 92.1) | 0.69 (0.68 - 0.70) | 0.65 (0.64 - 0.66) | 0.84 (0.83 - 0.86) |
| Primary and nursery education teaching professionals | 187,791 | 173,976 | 92.6 (92.5 - 92.8) | 92.8 (92.4 - 93.2) | 0.59 (0.58 - 0.60) | 0.59 (0.58 - 0.60) | 0.79 (0.77 - 0.80) |
| Special needs education teaching professionals | 14,427 | 13,323 | 92.3 (91.9 - 92.8) | 91.6 (90.0 - 93.2) | 0.62 (0.58 - 0.66) | 0.72 (0.67 - 0.76) | 0.91 (0.86 - 0.97) |
| Senior professionals of educational establishments | 65,028 | 60,875 | 93.6 (93.4 - 93.8) | 92.8 (92.0 - 93.6) | 0.51 (0.49 - 0.53) | 0.59 (0.57 - 0.61) | 0.76 (0.73 - 0.78) |
| Education advisers and school inspectors | 7,470 | 6,903 | 92.4 (91.8 - 93.0) | 91.4 (89.1 - 93.6) | 0.62 (0.57 - 0.67) | 0.71 (0.65 - 0.78) | 0.89 (0.81 - 0.97) |
| Teaching and other educational professionals n.e.c. | 57,925 | 50,517 | 87.2 (86.9 - 87.5) | 86.8 (86.0 - 87.6) | 1.10 (1.07 - 1.13) | 1.18 (1.15 - 1.21) | 1.34 (1.31 - 1.38) |
| Barristers and judges | 11,748 | 10,883 | 92.6 (92.2 - 93.1) | 92.5 (90.8 - 94.3) | 0.60 (0.56 - 0.64) | 0.58 (0.54 - 0.62) | 0.66 (0.61 - 0.70) |
| Solicitors | 59,739 | 55,615 | 93.1 (92.9 - 93.3) | 93.5 (92.7 - 94.3) | 0.55 (0.54 - 0.57) | 0.49 (0.47 - 0.50) | 0.60 (0.58 - 0.62) |
| Legal professionals n.e.c. | 21,500 | 18,857 | 87.7 (87.3 - 88.1) | 88.1 (86.8 - 89.3) | 1.05 (1.01 - 1.09) | 0.98 (0.94 - 1.02) | 0.95 (0.91 - 0.99) |
| Chartered and certified accountants | 116,527 | 106,523 | 91.4 (91.3 - 91.6) | 91.8 (91.3 - 92.4) | 0.70 (0.69 - 0.72) | 0.63 (0.61 - 0.64) | 0.75 (0.73 - 0.77) |
| Management consultants and business analysts | 65,406 | 58,241 | 89.0 (88.8 - 89.3) | 89.1 (88.4 - 89.8) | 0.92 (0.90 - 0.94) | 0.84 (0.82 - 0.86) | 0.95 (0.92 - 0.97) |
| Business and financial project management professionals | 87,039 | 79,094 | 90.9 (90.7 - 91.1) | 90.8 (90.2 - 91.5) | 0.75 (0.73 - 0.77) | 0.70 (0.69 - 0.72) | 0.82 (0.80 - 0.84) |
| Actuaries, economists and statisticians | 10,244 | 8,995 | 87.8 (87.2 - 88.4) | 88.9 (86.9 - 90.8) | 1.04 (0.98 - 1.10) | 0.81 (0.76 - 0.86) | 0.85 (0.80 - 0.90) |
| Business and related research professionals | 23,138 | 20,120 | 87.0 (86.5 - 87.4) | 88.1 (86.8 - 89.4) | 1.12 (1.08 - 1.17) | 0.96 (0.92 - 1.00) | 0.99 (0.95 - 1.03) |
| Business, research and administrative professionals n.e.c. | 19,177 | 17,632 | 91.9 (91.6 - 92.3) | 91.6 (90.2 - 92.9) | 0.66 (0.62 - 0.69) | 0.70 (0.66 - 0.74) | 0.85 (0.81 - 0.90) |
| Architects | 24,081 | 21,142 | 87.8 (87.4 - 88.2) | 88.1 (86.9 - 89.3) | 1.04 (1.00 - 1.08) | 0.92 (0.88 - 0.95) | 0.95 (0.91 - 0.99) |
| Town planning officers | 5,313 | 4,989 | 93.9 (93.3 - 94.5) | 94.3 (91.6 - 97.0) | 0.49 (0.43 - 0.54) | 0.41 (0.37 - 0.46) | 0.57 (0.50 - 0.63) |
| Quantity surveyors | 14,191 | 12,940 | 91.2 (90.7 - 91.7) | 91.6 (90.0 - 93.2) | 0.72 (0.68 - 0.77) | 0.59 (0.56 - 0.63) | 0.80 (0.75 - 0.85) |
| Chartered surveyors | 33,252 | 30,967 | 93.1 (92.9 - 93.4) | 92.8 (91.7 - 93.8) | 0.55 (0.53 - 0.58) | 0.51 (0.49 - 0.53) | 0.72 (0.68 - 0.75) |
| Chartered architectural technologists | 1,464 | 1,341 | 91.6 (90.2 - 93.0) | 91.9 (86.9 - 96.8) | 0.69 (0.57 - 0.83) | 0.57 (0.47 - 0.68) | 0.80 (0.66 - 0.97) |
| Construction project managers and related professionals | 29,960 | 27,689 | 92.4 (92.1 - 92.7) | 91.7 (90.6 - 92.8) | 0.61 (0.59 - 0.64) | 0.57 (0.55 - 0.60) | 0.73 (0.70 - 0.76) |
| Social workers | 42,730 | 37,826 | 88.5 (88.2 - 88.8) | 88.1 (87.2 - 89.0) | 0.97 (0.94 - 1.00) | 1.09 (1.05 - 1.12) | 1.06 (1.03 - 1.09) |
| Probation officers | 5,601 | 5,084 | 90.8 (90.0 - 91.5) | 90.7 (88.2 - 93.2) | 0.76 (0.70 - 0.83) | 0.75 (0.69 - 0.82) | 0.86 (0.78 - 0.94) |
| Clergy | 14,189 | 12,398 | 87.4 (86.8 - 87.9) | 85.6 (84.0 - 87.3) | 1.08 (1.03 - 1.14) | 1.19 (1.13 - 1.25) | 1.18 (1.12 - 1.24) |
| Welfare professionals n.e.c. | 14,644 | 12,647 | 86.4 (85.8 - 86.9) | 86.3 (84.7 - 87.8) | 1.18 (1.13 - 1.24) | 1.21 (1.16 - 1.27) | 1.30 (1.24 - 1.37) |
| Librarians | 9,870 | 9,215 | 93.4 (92.9 - 93.9) | 92.6 (90.6 - 94.6) | 0.53 (0.49 - 0.58) | 0.64 (0.59 - 0.69) | 0.74 (0.68 - 0.80) |
| Archivists and curators | 5,412 | 4,965 | 91.7 (91.0 - 92.5) | 91.8 (89.2 - 94.3) | 0.67 (0.61 - 0.74) | 0.66 (0.60 - 0.73) | 0.77 (0.70 - 0.85) |
| Quality control and planning engineers | 12,178 | 11,196 | 91.9 (91.5 - 92.4) | 91.6 (89.9 - 93.3) | 0.66 (0.62 - 0.70) | 0.61 (0.57 - 0.65) | 0.77 (0.72 - 0.82) |
| Quality assurance and regulatory professionals | 26,766 | 24,822 | 92.7 (92.4 - 93.0) | 92.3 (91.1 - 93.4) | 0.59 (0.56 - 0.61) | 0.58 (0.55 - 0.61) | 0.71 (0.68 - 0.74) |
| Environmental health professionals | 3,806 | 3,539 | 93.0 (92.2 - 93.8) | 92.8 (89.7 - 95.9) | 0.57 (0.50 - 0.64) | 0.58 (0.51 - 0.65) | 0.75 (0.66 - 0.86) |
| Journalists, newspaper and periodical editors | 34,236 | 30,980 | 90.5 (90.2 - 90.8) | 90.7 (89.7 - 91.7) | 0.79 (0.76 - 0.82) | 0.71 (0.69 - 0.74) | 0.81 (0.78 - 0.84) |
| Public relations professionals | 18,571 | 16,713 | 90.0 (89.6 - 90.4) | 90.8 (89.4 - 92.3) | 0.83 (0.79 - 0.87) | 0.73 (0.69 - 0.76) | 0.83 (0.79 - 0.87) |
| Advertising accounts managers and creative directors | 19,087 | 16,788 | 88.0 (87.5 - 88.4) | 89.1 (87.7 - 90.6) | 1.03 (0.98 - 1.07) | 0.84 (0.81 - 0.88) | 0.96 (0.92 - 1.01) |
| Laboratory technicians | 23,775 | 21,353 | 89.8 (89.4 - 90.2) | 89.5 (88.2 - 90.7) | 0.85 (0.82 - 0.89) | 0.85 (0.82 - 0.89) | 0.92 (0.88 - 0.96) |
| Electrical and electronics technicians | 6,134 | 5,457 | 89.0 (88.2 - 89.7) | 88.0 (85.6 - 90.3) | 0.93 (0.86 - 1.01) | 0.88 (0.81 - 0.96) | 1.04 (0.96 - 1.13) |
| Engineering technicians | 16,563 | 15,006 | 90.6 (90.2 - 91.0) | 90.1 (88.6 - 91.5) | 0.78 (0.74 - 0.82) | 0.71 (0.67 - 0.75) | 0.92 (0.87 - 0.97) |
| Building and civil engineering technicians | 4,081 | 3,661 | 89.7 (88.8 - 90.6) | 89.3 (86.4 - 92.2) | 0.86 (0.78 - 0.95) | 0.81 (0.73 - 0.89) | 0.92 (0.83 - 1.03) |
| Quality assurance technicians | 9,921 | 8,994 | 90.7 (90.1 - 91.2) | 90.7 (88.8 - 92.5) | 0.77 (0.72 - 0.83) | 0.72 (0.67 - 0.77) | 0.81 (0.76 - 0.87) |
| Planning, process and production technicians | 10,955 | 10,067 | 91.9 (91.4 - 92.4) | 91.5 (89.7 - 93.3) | 0.66 (0.62 - 0.71) | 0.62 (0.58 - 0.66) | 0.74 (0.69 - 0.79) |
| Science, engineering and production technicians n.e.c. | 40,041 | 35,883 | 89.6 (89.3 - 89.9) | 88.7 (87.8 - 89.6) | 0.87 (0.84 - 0.90) | 0.85 (0.82 - 0.88) | 0.96 (0.93 - 0.99) |
| Architectural and town planning technicians | 6,338 | 5,572 | 87.9 (87.1 - 88.7) | 89.6 (87.1 - 92.0) | 1.03 (0.96 - 1.11) | 0.81 (0.75 - 0.87) | 0.88 (0.81 - 0.95) |
| Draughtspersons | 14,241 | 12,824 | 90.0 (89.6 - 90.5) | 89.8 (88.3 - 91.4) | 0.83 (0.78 - 0.87) | 0.75 (0.71 - 0.79) | 0.92 (0.87 - 0.97) |
| IT operations technicians | 46,473 | 40,997 | 88.2 (87.9 - 88.5) | 88.7 (87.9 - 89.6) | 1.00 (0.97 - 1.03) | 0.85 (0.83 - 0.88) | 0.93 (0.90 - 0.95) |
| IT user support technicians | 40,089 | 35,500 | 88.6 (88.2 - 88.9) | 89.5 (88.5 - 90.4) | 0.97 (0.94 - 1.00) | 0.78 (0.75 - 0.80) | 0.85 (0.82 - 0.87) |
| Paramedics | 9,873 | 9,242 | 93.6 (93.1 - 94.1) | 93.7 (91.8 - 95.6) | 0.51 (0.47 - 0.55) | 0.46 (0.42 - 0.50) | 0.73 (0.67 - 0.79) |
| Dispensing opticians | 2,498 | 2,321 | 92.9 (91.9 - 93.9) | 93.1 (89.3 - 96.9) | 0.57 (0.49 - 0.67) | 0.55 (0.47 - 0.64) | 0.69 (0.59 - 0.81) |
| Pharmaceutical technicians | 10,991 | 10,140 | 92.3 (91.8 - 92.8) | 92.2 (90.4 - 94.0) | 0.63 (0.59 - 0.67) | 0.66 (0.61 - 0.71) | 0.79 (0.74 - 0.85) |
| Medical and dental technicians | 12,034 | 10,895 | 90.5 (90.0 - 91.1) | 90.1 (88.4 - 91.8) | 0.78 (0.74 - 0.83) | 0.80 (0.75 - 0.85) | 0.91 (0.85 - 0.97) |
| Health associate professionals n.e.c. | 21,136 | 16,318 | 77.2 (76.6 - 77.8) | 76.7 (75.5 - 77.9) | 2.22 (2.15 - 2.29) | 2.60 (2.51 - 2.68) | 3.00 (2.90 - 3.10) |
| Youth and community workers | 25,724 | 22,235 | 86.4 (86.0 - 86.9) | 86.4 (85.2 - 87.5) | 1.18 (1.14 - 1.22) | 1.22 (1.17 - 1.26) | 1.23 (1.18 - 1.28) |
| Child and early years officers | 5,609 | 5,118 | 91.2 (90.5 - 92.0) | 90.5 (87.9 - 93.0) | 0.72 (0.66 - 0.79) | 0.83 (0.76 - 0.91) | 0.92 (0.84 - 1.02) |
| Housing officers | 21,840 | 18,925 | 86.7 (86.2 - 87.1) | 86.2 (85.0 - 87.5) | 1.15 (1.11 - 1.20) | 1.23 (1.19 - 1.28) | 1.12 (1.07 - 1.16) |
| Counsellors | 11,491 | 9,842 | 85.6 (85.0 - 86.3) | 84.6 (82.9 - 86.4) | 1.26 (1.19 - 1.32) | 1.49 (1.41 - 1.57) | 1.66 (1.57 - 1.75) |
| Welfare and housing associate professionals n.e.c. | 57,200 | 49,903 | 87.2 (87.0 - 87.5) | 86.7 (86.0 - 87.5) | 1.10 (1.07 - 1.12) | 1.20 (1.17 - 1.23) | 1.21 (1.18 - 1.24) |
| NCOs and other ranks | 13,981 | 11,860 | 84.8 (84.2 - 85.4) | 84.9 (83.3 - 86.4) | 1.34 (1.28 - 1.40) | 1.16 (1.11 - 1.22) | 1.26 (1.20 - 1.32) |
| Police officers (sergeant and below) | 108,003 | 101,946 | 94.4 (94.3 - 94.5) | 94.4 (93.8 - 95.0) | 0.44 (0.43 - 0.46) | 0.39 (0.38 - 0.40) | 0.53 (0.51 - 0.54) |
| Fire service officers (watch manager and below) | 24,700 | 22,331 | 90.4 (90.0 - 90.8) | 90.1 (88.9 - 91.2) | 0.79 (0.76 - 0.83) | 0.71 (0.68 - 0.74) | 0.93 (0.89 - 0.97) |
| Prison service officers (below principal officer) | 22,562 | 20,605 | 91.3 (91.0 - 91.7) | 90.5 (89.3 - 91.8) | 0.71 (0.68 - 0.75) | 0.72 (0.68 - 0.75) | 0.81 (0.77 - 0.85) |
| Police community support officers | 7,452 | 6,835 | 91.7 (91.1 - 92.3) | 91.8 (89.7 - 94.0) | 0.68 (0.62 - 0.73) | 0.62 (0.57 - 0.67) | 0.66 (0.61 - 0.72) |
| Protective service associate professionals n.e.c. | 16,061 | 14,507 | 90.3 (89.9 - 90.8) | 90.0 (88.6 - 91.5) | 0.80 (0.76 - 0.85) | 0.76 (0.72 - 0.80) | 0.82 (0.78 - 0.87) |
| Artists | 20,821 | 17,174 | 82.5 (82.0 - 83.0) | 82.1 (80.9 - 83.4) | 1.59 (1.54 - 1.65) | 1.61 (1.55 - 1.67) | 1.83 (1.77 - 1.90) |
| Authors, writers and translators | 28,632 | 24,145 | 84.3 (83.9 - 84.7) | 84.2 (83.1 - 85.3) | 1.39 (1.35 - 1.44) | 1.41 (1.36 - 1.45) | 1.30 (1.25 - 1.34) |
| Actors, entertainers and presenters | 19,150 | 15,422 | 80.5 (80.0 - 81.1) | 81.0 (79.7 - 82.3) | 1.81 (1.75 - 1.88) | 1.66 (1.60 - 1.72) | 1.61 (1.55 - 1.68) |
| Dancers and choreographers | 6,342 | 5,275 | 83.2 (82.3 - 84.1) | 84.2 (81.9 - 86.6) | 1.52 (1.42 - 1.62) | 1.46 (1.37 - 1.56) | 1.60 (1.49 - 1.72) |
| Musicians | 15,755 | 13,066 | 82.9 (82.3 - 83.5) | 83.0 (81.5 - 84.4) | 1.54 (1.48 - 1.61) | 1.41 (1.35 - 1.47) | 1.48 (1.42 - 1.55) |
| Arts officers, producers and directors | 33,255 | 29,002 | 87.2 (86.9 - 87.6) | 87.7 (86.7 - 88.8) | 1.10 (1.06 - 1.14) | 0.95 (0.92 - 0.99) | 1.01 (0.98 - 1.04) |
| Photographers, audio-visual and broadcasting equipment operators | 30,398 | 25,869 | 85.1 (84.7 - 85.5) | 85.2 (84.2 - 86.2) | 1.31 (1.27 - 1.36) | 1.18 (1.14 - 1.22) | 1.26 (1.22 - 1.31) |
| Graphic designers | 38,700 | 33,977 | 87.8 (87.5 - 88.1) | 88.6 (87.6 - 89.6) | 1.04 (1.01 - 1.07) | 0.86 (0.83 - 0.89) | 1.02 (0.99 - 1.06) |
| Product, clothing and related designers | 30,806 | 26,549 | 86.2 (85.8 - 86.6) | 86.3 (85.3 - 87.4) | 1.20 (1.16 - 1.24) | 1.16 (1.12 - 1.20) | 1.26 (1.22 - 1.30) |
| Sports players | 4,217 | 3,465 | 82.2 (81.0 - 83.3) | 84.4 (81.4 - 87.4) | 1.63 (1.50 - 1.76) | 1.22 (1.13 - 1.33) | 1.41 (1.30 - 1.53) |
| Sports coaches, instructors and officials | 21,203 | 18,384 | 86.7 (86.2 - 87.2) | 86.9 (85.6 - 88.2) | 1.15 (1.11 - 1.20) | 1.08 (1.03 - 1.12) | 1.30 (1.25 - 1.35) |
| Fitness instructors | 18,657 | 14,511 | 77.8 (77.2 - 78.4) | 78.6 (77.3 - 79.9) | 2.14 (2.07 - 2.22) | 2.06 (1.99 - 2.13) | 2.29 (2.21 - 2.38) |
| Air traffic controllers | 1,694 | 1,560 | 92.1 (90.8 - 93.4) | 92.0 (87.5 - 96.6) | 0.64 (0.54 - 0.77) | 0.58 (0.48 - 0.69) | 0.86 (0.72 - 1.03) |
| Aircraft pilots and flight engineers | 8,024 | 7,093 | 88.4 (87.7 - 89.1) | 87.8 (85.8 - 89.9) | 0.98 (0.92 - 1.05) | 0.92 (0.86 - 0.99) | 1.37 (1.28 - 1.47) |
| Ship and hovercraft officers | 3,756 | 3,291 | 87.6 (86.6 - 88.7) | 85.8 (82.7 - 89.0) | 1.06 (0.96 - 1.17) | 1.05 (0.95 - 1.15) | 1.42 (1.28 - 1.57) |
| Legal associate professionals | 26,646 | 24,104 | 90.5 (90.1 - 90.8) | 90.8 (89.6 - 91.9) | 0.79 (0.76 - 0.82) | 0.75 (0.72 - 0.78) | 0.85 (0.81 - 0.89) |
| Estimators, valuers and assessors | 26,441 | 24,275 | 91.8 (91.5 - 92.1) | 91.5 (90.3 - 92.6) | 0.67 (0.64 - 0.70) | 0.64 (0.61 - 0.67) | 0.81 (0.77 - 0.84) |
| Brokers | 29,462 | 26,112 | 88.6 (88.3 - 89.0) | 88.8 (87.7 - 89.9) | 0.96 (0.93 - 1.00) | 0.85 (0.82 - 0.88) | 0.94 (0.91 - 0.98) |
| Insurance underwriters | 14,945 | 13,966 | 93.4 (93.1 - 93.8) | 93.8 (92.3 - 95.4) | 0.53 (0.49 - 0.56) | 0.47 (0.44 - 0.50) | 0.60 (0.56 - 0.64) |
| Finance and investment analysts and advisers | 80,758 | 72,319 | 89.6 (89.3 - 89.8) | 90.1 (89.5 - 90.8) | 0.87 (0.85 - 0.89) | 0.76 (0.74 - 0.78) | 0.85 (0.83 - 0.87) |
| Taxation experts | 14,574 | 13,652 | 93.7 (93.3 - 94.1) | 93.6 (92.0 - 95.2) | 0.51 (0.47 - 0.54) | 0.48 (0.45 - 0.52) | 0.62 (0.58 - 0.66) |
| Importers and exporters | 3,835 | 3,294 | 85.9 (84.8 - 87.0) | 85.8 (82.9 - 88.7) | 1.23 (1.12 - 1.35) | 1.18 (1.08 - 1.29) | 1.05 (0.96 - 1.16) |
| Financial and accounting technicians | 7,926 | 7,267 | 91.7 (91.1 - 92.3) | 91.8 (89.6 - 93.9) | 0.68 (0.63 - 0.74) | 0.66 (0.61 - 0.72) | 0.72 (0.66 - 0.78) |
| Financial accounts managers | 59,742 | 54,794 | 91.7 (91.5 - 91.9) | 91.8 (91.1 - 92.6) | 0.68 (0.66 - 0.70) | 0.63 (0.61 - 0.65) | 0.73 (0.71 - 0.76) |
| Business and related associate professionals n.e.c. | 46,422 | 41,696 | 89.8 (89.5 - 90.1) | 90.4 (89.5 - 91.3) | 0.85 (0.82 - 0.88) | 0.76 (0.74 - 0.78) | 0.83 (0.80 - 0.86) |
| Buyers and procurement officers | 29,858 | 27,363 | 91.6 (91.3 - 92.0) | 91.9 (90.8 - 93.0) | 0.68 (0.66 - 0.71) | 0.63 (0.61 - 0.66) | 0.75 (0.71 - 0.78) |
| Business sales executives | 82,738 | 73,409 | 88.7 (88.5 - 88.9) | 88.9 (88.3 - 89.6) | 0.95 (0.93 - 0.97) | 0.86 (0.84 - 0.88) | 0.95 (0.93 - 0.97) |
| Marketing associate professionals | 46,756 | 41,351 | 88.4 (88.2 - 88.7) | 89.3 (88.4 - 90.2) | 0.98 (0.95 - 1.01) | 0.86 (0.84 - 0.88) | 0.96 (0.93 - 0.98) |
| Estate agents and auctioneers | 27,753 | 24,626 | 88.7 (88.4 - 89.1) | 88.7 (87.6 - 89.8) | 0.95 (0.92 - 0.99) | 0.92 (0.88 - 0.95) | 1.01 (0.98 - 1.05) |
| Sales accounts and business development managers | 190,004 | 173,570 | 91.4 (91.2 - 91.5) | 91.6 (91.2 - 92.1) | 0.71 (0.70 - 0.72) | 0.63 (0.62 - 0.64) | 0.77 (0.76 - 0.78) |
| Conference and exhibition managers and organisers | 22,076 | 19,403 | 87.9 (87.5 - 88.3) | 88.9 (87.6 - 90.2) | 1.03 (0.99 - 1.08) | 0.92 (0.88 - 0.96) | 0.94 (0.90 - 0.98) |
| Conservation and environmental associate professionals | 3,277 | 2,947 | 89.9 (88.9 - 91.0) | 89.6 (86.3 - 92.9) | 0.84 (0.75 - 0.94) | 0.79 (0.71 - 0.89) | 1.01 (0.90 - 1.14) |
| Public services associate professionals | 23,308 | 21,478 | 92.1 (91.8 - 92.5) | 91.5 (90.3 - 92.8) | 0.64 (0.61 - 0.67) | 0.71 (0.67 - 0.74) | 0.74 (0.71 - 0.78) |
| Human resources and industrial relations officers | 69,794 | 62,547 | 89.6 (89.4 - 89.8) | 90.6 (89.9 - 91.3) | 0.87 (0.85 - 0.89) | 0.77 (0.75 - 0.79) | 0.88 (0.86 - 0.91) |
| Vocational and industrial trainers and instructors | 71,845 | 65,206 | 90.8 (90.5 - 91.0) | 90.2 (89.5 - 90.9) | 0.76 (0.74 - 0.78) | 0.80 (0.78 - 0.82) | 0.97 (0.95 - 1.00) |
| Careers advisers and vocational guidance specialists | 10,454 | 9,361 | 89.5 (89.0 - 90.1) | 89.3 (87.5 - 91.1) | 0.88 (0.82 - 0.93) | 0.92 (0.86 - 0.98) | 1.05 (0.99 - 1.12) |
| Inspectors of standards and regulations | 9,981 | 9,269 | 92.9 (92.4 - 93.4) | 92.2 (90.2 - 94.1) | 0.58 (0.53 - 0.62) | 0.59 (0.55 - 0.64) | 0.74 (0.68 - 0.80) |
| Health and safety officers | 17,253 | 15,931 | 92.3 (91.9 - 92.7) | 91.5 (90.0 - 93.0) | 0.62 (0.59 - 0.66) | 0.64 (0.60 - 0.67) | 0.81 (0.77 - 0.86) |
| National government administrative occupations | 150,201 | 138,324 | 92.1 (92.0 - 92.2) | 91.4 (90.9 - 91.9) | 0.64 (0.63 - 0.65) | 0.70 (0.69 - 0.71) | 0.75 (0.74 - 0.76) |
| Local government administrative occupations | 73,705 | 67,995 | 92.3 (92.1 - 92.4) | 91.5 (90.8 - 92.2) | 0.63 (0.61 - 0.65) | 0.71 (0.69 - 0.72) | 0.76 (0.74 - 0.78) |
| Officers of non-governmental organisations | 12,062 | 10,942 | 90.7 (90.2 - 91.2) | 90.1 (88.4 - 91.8) | 0.77 (0.72 - 0.82) | 0.85 (0.80 - 0.91) | 0.88 (0.83 - 0.94) |
| Credit controllers | 22,476 | 20,352 | 90.5 (90.2 - 90.9) | 90.5 (89.2 - 91.7) | 0.78 (0.75 - 0.82) | 0.81 (0.77 - 0.84) | 0.82 (0.78 - 0.85) |
| Book-keepers, payroll managers and wages clerks | 159,886 | 145,547 | 91.0 (90.9 - 91.2) | 90.6 (90.1 - 91.0) | 0.74 (0.72 - 0.75) | 0.80 (0.78 - 0.81) | 0.83 (0.81 - 0.84) |
| Bank and post office clerks | 94,616 | 86,420 | 91.3 (91.2 - 91.5) | 90.7 (90.1 - 91.3) | 0.71 (0.69 - 0.73) | 0.76 (0.75 - 0.78) | 0.77 (0.75 - 0.79) |
| Finance officers | 15,058 | 13,743 | 91.3 (90.8 - 91.7) | 90.3 (88.8 - 91.9) | 0.72 (0.68 - 0.76) | 0.80 (0.75 - 0.84) | 0.83 (0.78 - 0.88) |
| Financial administrative occupations n.e.c. | 64,172 | 57,389 | 89.4 (89.2 - 89.7) | 89.3 (88.5 - 90.0) | 0.89 (0.86 - 0.91) | 0.92 (0.90 - 0.94) | 0.88 (0.86 - 0.90) |
| Records clerks and assistants | 56,035 | 51,176 | 91.3 (91.1 - 91.6) | 90.8 (90.0 - 91.6) | 0.71 (0.69 - 0.73) | 0.77 (0.75 - 0.79) | 0.80 (0.77 - 0.82) |
| Pensions and insurance clerks and assistants | 41,863 | 38,273 | 91.4 (91.2 - 91.7) | 91.7 (90.8 - 92.6) | 0.70 (0.68 - 0.73) | 0.67 (0.65 - 0.70) | 0.77 (0.74 - 0.80) |
| Stock control clerks and assistants | 34,036 | 30,437 | 89.4 (89.1 - 89.8) | 89.1 (88.1 - 90.1) | 0.89 (0.86 - 0.92) | 0.85 (0.83 - 0.88) | 0.86 (0.83 - 0.89) |
| Transport and distribution clerks and assistants | 30,535 | 27,327 | 89.5 (89.2 - 89.8) | 89.4 (88.3 - 90.4) | 0.88 (0.85 - 0.91) | 0.83 (0.80 - 0.86) | 0.83 (0.80 - 0.87) |
| Library clerks and assistants | 15,067 | 13,834 | 91.8 (91.4 - 92.3) | 90.7 (89.1 - 92.3) | 0.67 (0.63 - 0.71) | 0.79 (0.74 - 0.84) | 0.85 (0.80 - 0.90) |
| Human resources administrative occupations | 18,961 | 17,144 | 90.4 (90.0 - 90.8) | 90.7 (89.3 - 92.0) | 0.79 (0.76 - 0.83) | 0.81 (0.77 - 0.85) | 0.85 (0.81 - 0.89) |
| Sales administrators | 34,644 | 31,213 | 90.1 (89.8 - 90.4) | 90.5 (89.5 - 91.5) | 0.82 (0.80 - 0.85) | 0.81 (0.78 - 0.84) | 0.89 (0.86 - 0.92) |
| Other administrative occupations n.e.c. | 320,894 | 288,971 | 90.1 (89.9 - 90.2) | 89.5 (89.2 - 89.8) | 0.82 (0.82 - 0.83) | 0.91 (0.90 - 0.92) | 0.92 (0.91 - 0.93) |
| Office managers | 85,259 | 78,780 | 92.4 (92.2 - 92.6) | 91.8 (91.2 - 92.5) | 0.62 (0.60 - 0.63) | 0.68 (0.66 - 0.69) | 0.76 (0.74 - 0.78) |
| Office supervisors | 17,917 | 16,574 | 92.5 (92.1 - 92.9) | 92.4 (91.0 - 93.8) | 0.61 (0.57 - 0.64) | 0.61 (0.58 - 0.65) | 0.66 (0.62 - 0.70) |
| Medical secretaries | 33,188 | 31,002 | 93.4 (93.1 - 93.7) | 92.4 (91.3 - 93.6) | 0.53 (0.51 - 0.55) | 0.67 (0.64 - 0.70) | 0.74 (0.71 - 0.77) |
| Legal secretaries | 30,679 | 28,001 | 91.3 (91.0 - 91.6) | 90.9 (89.8 - 92.0) | 0.72 (0.69 - 0.75) | 0.82 (0.78 - 0.85) | 0.85 (0.81 - 0.88) |
| School secretaries | 34,524 | 32,739 | 94.8 (94.6 - 95.1) | 92.8 (91.6 - 94.0) | 0.41 (0.39 - 0.43) | 0.54 (0.52 - 0.57) | 0.62 (0.59 - 0.65) |
| Company secretaries | 23,868 | 22,075 | 92.5 (92.2 - 92.8) | 90.9 (89.5 - 92.3) | 0.61 (0.58 - 0.64) | 0.79 (0.76 - 0.83) | 0.86 (0.82 - 0.91) |
| Personal assistants and other secretaries | 175,722 | 158,881 | 90.4 (90.3 - 90.6) | 89.6 (89.2 - 90.1) | 0.79 (0.78 - 0.81) | 0.93 (0.91 - 0.94) | 0.94 (0.92 - 0.96) |
| Receptionists | 118,749 | 105,211 | 88.6 (88.4 - 88.8) | 87.5 (86.9 - 88.0) | 0.96 (0.95 - 0.98) | 1.12 (1.10 - 1.14) | 1.06 (1.04 - 1.08) |
| Typists and related keyboard occupations | 30,168 | 26,422 | 87.6 (87.2 - 88.0) | 87.0 (85.9 - 88.0) | 1.06 (1.03 - 1.10) | 1.14 (1.10 - 1.18) | 1.08 (1.05 - 1.12) |
| Farmers | 38,490 | 35,756 | 92.9 (92.6 - 93.2) | 91.2 (90.2 - 92.3) | 0.57 (0.55 - 0.60) | 0.61 (0.59 - 0.64) | 0.76 (0.73 - 0.79) |
| Horticultural trades | 8,101 | 7,101 | 87.7 (86.9 - 88.4) | 86.5 (84.5 - 88.6) | 1.06 (0.99 - 1.13) | 1.14 (1.06 - 1.22) | 1.15 (1.07 - 1.23) |
| Gardeners and landscape gardeners | 66,824 | 57,193 | 85.6 (85.3 - 85.9) | 83.8 (83.1 - 84.5) | 1.26 (1.24 - 1.29) | 1.26 (1.23 - 1.29) | 1.33 (1.30 - 1.36) |
| Groundsmen and greenkeepers | 12,292 | 10,911 | 88.8 (88.2 - 89.3) | 88.0 (86.4 - 89.7) | 0.95 (0.90 - 1.00) | 0.86 (0.81 - 0.91) | 0.97 (0.92 - 1.03) |
| Agricultural and fishing trades n.e.c. | 7,923 | 6,596 | 83.3 (82.4 - 84.1) | 83.2 (81.1 - 85.2) | 1.51 (1.42 - 1.60) | 1.31 (1.24 - 1.39) | 1.58 (1.49 - 1.68) |
| Smiths and forge workers | 2,432 | 2,110 | 86.8 (85.4 - 88.1) | 85.5 (81.8 - 89.3) | 1.14 (1.02 - 1.29) | 1.08 (0.96 - 1.22) | 1.31 (1.16 - 1.48) |
| Moulders, core makers and die casters | 1,344 | 1,163 | 86.5 (84.7 - 88.4) | 83.7 (78.5 - 88.9) | 1.17 (1.00 - 1.36) | 1.20 (1.02 - 1.40) | 1.04 (0.89 - 1.23) |
| Sheet metal workers | 8,282 | 7,389 | 89.2 (88.5 - 89.9) | 87.3 (85.2 - 89.3) | 0.91 (0.85 - 0.97) | 0.89 (0.83 - 0.95) | 0.97 (0.90 - 1.04) |
| Metal plate workers, and riveters | 3,129 | 2,801 | 89.5 (88.4 - 90.6) | 86.2 (82.6 - 89.8) | 0.88 (0.78 - 0.98) | 0.93 (0.83 - 1.05) | 1.09 (0.97 - 1.22) |
| Welding trades | 30,831 | 25,965 | 84.2 (83.8 - 84.6) | 82.6 (81.6 - 83.6) | 1.41 (1.36 - 1.45) | 1.32 (1.28 - 1.37) | 1.30 (1.26 - 1.35) |
| Pipe fitters | 5,263 | 4,552 | 86.5 (85.6 - 87.4) | 84.5 (82.0 - 87.1) | 1.17 (1.08 - 1.27) | 1.11 (1.02 - 1.20) | 1.25 (1.15 - 1.35) |
| Metal machining setters and setter-operators | 20,704 | 18,368 | 88.7 (88.3 - 89.1) | 86.7 (85.4 - 88.0) | 0.95 (0.91 - 1.00) | 0.93 (0.89 - 0.97) | 0.96 (0.92 - 1.00) |
| Tool makers, tool fitters and markers-out | 5,733 | 5,271 | 91.9 (91.2 - 92.6) | 89.7 (87.0 - 92.3) | 0.66 (0.60 - 0.72) | 0.68 (0.62 - 0.75) | 0.82 (0.74 - 0.90) |
| Metal working production and maintenance fitters | 96,624 | 85,667 | 88.7 (88.5 - 88.9) | 86.9 (86.3 - 87.5) | 0.96 (0.94 - 0.98) | 0.93 (0.91 - 0.94) | 1.02 (1.00 - 1.05) |
| Precision instrument makers and repairers | 7,637 | 6,828 | 89.4 (88.7 - 90.1) | 88.3 (86.1 - 90.4) | 0.89 (0.83 - 0.96) | 0.85 (0.79 - 0.92) | 1.04 (0.97 - 1.13) |
| Air-conditioning and refrigeration engineers | 6,128 | 5,192 | 84.7 (83.8 - 85.6) | 85.4 (83.1 - 87.8) | 1.35 (1.26 - 1.45) | 1.10 (1.03 - 1.18) | 1.24 (1.15 - 1.33) |
| Vehicle technicians, mechanics and electricians | 83,650 | 70,993 | 84.9 (84.6 - 85.1) | 84.4 (83.8 - 85.0) | 1.34 (1.31 - 1.36) | 1.18 (1.16 - 1.21) | 1.25 (1.22 - 1.27) |
| Vehicle body builders and repairers | 13,259 | 11,323 | 85.4 (84.8 - 86.0) | 84.3 (82.8 - 85.9) | 1.28 (1.22 - 1.35) | 1.18 (1.12 - 1.24) | 1.23 (1.17 - 1.29) |
| Vehicle paint technicians | 7,785 | 6,564 | 84.3 (83.5 - 85.1) | 84.1 (82.1 - 86.1) | 1.39 (1.31 - 1.48) | 1.21 (1.14 - 1.29) | 1.22 (1.15 - 1.30) |
| Aircraft maintenance and related trades | 10,096 | 9,233 | 91.5 (90.9 - 92.0) | 90.1 (88.1 - 92.1) | 0.70 (0.65 - 0.75) | 0.70 (0.65 - 0.75) | 0.92 (0.86 - 0.99) |
| Boat and ship builders and repairers | 5,196 | 4,547 | 87.5 (86.6 - 88.4) | 86.4 (83.8 - 89.0) | 1.07 (0.99 - 1.16) | 0.99 (0.92 - 1.08) | 1.19 (1.10 - 1.30) |
| Rail and rolling stock builders and repairers | 4,675 | 4,154 | 88.9 (88.0 - 89.8) | 87.7 (85.0 - 90.4) | 0.94 (0.86 - 1.03) | 0.88 (0.81 - 0.97) | 0.94 (0.86 - 1.03) |
| Electricians and electrical fitters | 89,939 | 77,887 | 86.6 (86.4 - 86.8) | 86.1 (85.5 - 86.7) | 1.16 (1.14 - 1.18) | 1.02 (1.00 - 1.04) | 1.20 (1.17 - 1.22) |
| Telecommunications engineers | 27,836 | 24,638 | 88.5 (88.1 - 88.9) | 86.9 (85.8 - 88.0) | 0.97 (0.94 - 1.01) | 0.94 (0.90 - 0.97) | 0.94 (0.91 - 0.98) |
| TV, video and audio engineers | 4,739 | 3,952 | 83.4 (82.3 - 84.5) | 83.4 (80.8 - 86.0) | 1.49 (1.38 - 1.61) | 1.29 (1.19 - 1.39) | 1.25 (1.16 - 1.36) |
| IT engineers | 17,936 | 15,167 | 84.6 (84.0 - 85.1) | 85.2 (83.8 - 86.6) | 1.37 (1.31 - 1.43) | 1.11 (1.06 - 1.16) | 1.11 (1.06 - 1.16) |
| Electrical and electronic trades n.e.c. | 35,943 | 31,968 | 88.9 (88.6 - 89.3) | 88.1 (87.1 - 89.1) | 0.93 (0.90 - 0.96) | 0.86 (0.83 - 0.89) | 0.96 (0.93 - 0.99) |
| Skilled metal, electrical and electronic trades supervisors | 26,298 | 24,312 | 92.4 (92.1 - 92.8) | 91.3 (90.1 - 92.5) | 0.61 (0.58 - 0.64) | 0.59 (0.56 - 0.62) | 0.72 (0.69 - 0.75) |
| Steel erectors | 4,781 | 4,079 | 85.3 (84.3 - 86.3) | 84.6 (82.0 - 87.2) | 1.29 (1.19 - 1.40) | 1.15 (1.06 - 1.25) | 1.23 (1.13 - 1.34) |
| Bricklayers and masons | 35,742 | 30,489 | 85.3 (84.9 - 85.7) | 83.4 (82.5 - 84.4) | 1.29 (1.26 - 1.33) | 1.23 (1.19 - 1.26) | 1.35 (1.31 - 1.39) |
| Roofers, roof tilers and slaters | 25,969 | 21,078 | 81.2 (80.7 - 81.6) | 80.3 (79.2 - 81.4) | 1.74 (1.69 - 1.80) | 1.56 (1.51 - 1.61) | 1.55 (1.50 - 1.60) |
| Plumbers and heating and ventilating engineers | 68,978 | 59,462 | 86.2 (85.9 - 86.5) | 85.3 (84.6 - 86.0) | 1.20 (1.18 - 1.23) | 1.08 (1.06 - 1.11) | 1.25 (1.22 - 1.28) |
| Carpenters and joiners | 110,235 | 92,977 | 84.3 (84.1 - 84.6) | 83.2 (82.7 - 83.7) | 1.40 (1.37 - 1.42) | 1.27 (1.25 - 1.30) | 1.30 (1.28 - 1.32) |
| Glaziers, window fabricators and fitters | 23,223 | 19,427 | 83.7 (83.2 - 84.1) | 83.3 (82.1 - 84.5) | 1.47 (1.42 - 1.52) | 1.28 (1.23 - 1.32) | 1.26 (1.22 - 1.31) |
| Construction and building trades n.e.c. | 130,834 | 108,984 | 83.3 (83.1 - 83.5) | 81.2 (80.7 - 81.7) | 1.51 (1.49 - 1.53) | 1.45 (1.42 - 1.47) | 1.29 (1.27 - 1.31) |
| Plasterers | 26,578 | 21,310 | 80.2 (79.7 - 80.7) | 79.4 (78.3 - 80.5) | 1.86 (1.80 - 1.91) | 1.66 (1.61 - 1.71) | 1.62 (1.57 - 1.67) |
| Floorers and wall tilers | 20,565 | 17,111 | 83.2 (82.7 - 83.7) | 82.5 (81.3 - 83.8) | 1.51 (1.46 - 1.57) | 1.35 (1.30 - 1.40) | 1.38 (1.33 - 1.43) |
| Painters and decorators | 71,662 | 58,697 | 81.9 (81.6 - 82.2) | 78.6 (77.9 - 79.3) | 1.66 (1.63 - 1.69) | 1.69 (1.66 - 1.72) | 1.43 (1.40 - 1.46) |
| Construction and building trades supervisors | 21,478 | 19,220 | 89.5 (89.1 - 89.9) | 87.5 (86.2 - 88.8) | 0.88 (0.84 - 0.92) | 0.86 (0.82 - 0.90) | 0.90 (0.86 - 0.94) |
| Weavers and knitters | 1,800 | 1,555 | 86.4 (84.8 - 88.0) | 83.0 (78.2 - 87.7) | 1.18 (1.03 - 1.35) | 1.36 (1.19 - 1.56) | 1.06 (0.92 - 1.21) |
| Upholsterers | 8,962 | 7,850 | 87.6 (86.9 - 88.3) | 85.1 (83.1 - 87.1) | 1.06 (1.00 - 1.13) | 1.14 (1.07 - 1.21) | 1.08 (1.01 - 1.15) |
| Footwear and leather working trades | 6,006 | 5,163 | 86.0 (85.1 - 86.8) | 83.3 (80.8 - 85.8) | 1.22 (1.14 - 1.32) | 1.39 (1.29 - 1.49) | 1.15 (1.06 - 1.24) |
| Tailors and dressmakers | 8,687 | 7,030 | 80.9 (80.1 - 81.8) | 79.7 (77.8 - 81.6) | 1.77 (1.68 - 1.86) | 1.98 (1.88 - 2.09) | 1.16 (1.09 - 1.22) |
| Textiles, garments and related trades n.e.c. | 2,943 | 2,527 | 85.9 (84.6 - 87.1) | 84.2 (80.8 - 87.6) | 1.23 (1.11 - 1.37) | 1.38 (1.24 - 1.53) | 1.16 (1.04 - 1.30) |
| Pre-press technicians | 2,854 | 2,553 | 89.5 (88.3 - 90.6) | 88.3 (84.8 - 91.8) | 0.88 (0.78 - 1.00) | 0.88 (0.78 - 0.99) | 0.97 (0.86 - 1.10) |
| Printers | 26,199 | 23,269 | 88.8 (88.4 - 89.2) | 87.8 (86.6 - 88.9) | 0.94 (0.91 - 0.98) | 0.90 (0.86 - 0.93) | 0.96 (0.92 - 1.00) |
| Print finishing and binding workers | 10,495 | 9,305 | 88.7 (88.1 - 89.3) | 87.3 (85.5 - 89.2) | 0.96 (0.90 - 1.02) | 0.98 (0.92 - 1.04) | 0.99 (0.93 - 1.05) |
| Butchers | 16,142 | 13,553 | 84.0 (83.4 - 84.5) | 82.2 (80.8 - 83.6) | 1.43 (1.37 - 1.49) | 1.38 (1.32 - 1.44) | 1.02 (0.98 - 1.07) |
| Bakers and flour confectioners | 14,325 | 12,429 | 86.8 (86.2 - 87.3) | 86.3 (84.8 - 87.9) | 1.14 (1.09 - 1.20) | 1.11 (1.06 - 1.17) | 0.94 (0.89 - 0.99) |
| Fishmongers and poultry dressers | 2,627 | 2,236 | 85.1 (83.8 - 86.5) | 83.4 (79.8 - 86.9) | 1.31 (1.18 - 1.46) | 1.32 (1.18 - 1.47) | 1.12 (1.00 - 1.25) |
| Chefs | 105,199 | 86,849 | 82.6 (82.3 - 82.8) | 83.2 (82.6 - 83.7) | 1.59 (1.57 - 1.62) | 1.37 (1.35 - 1.39) | 0.91 (0.90 - 0.93) |
| Cooks | 46,852 | 41,348 | 88.3 (88.0 - 88.5) | 86.1 (85.2 - 87.0) | 1.00 (0.97 - 1.03) | 1.20 (1.17 - 1.23) | 0.97 (0.94 - 1.00) |
| Catering and bar managers | 62,157 | 53,912 | 86.7 (86.5 - 87.0) | 85.6 (84.9 - 86.4) | 1.15 (1.12 - 1.17) | 1.23 (1.20 - 1.26) | 1.02 (1.00 - 1.05) |
| Glass and ceramics makers, decorators and finishers | 7,627 | 6,756 | 88.6 (87.9 - 89.3) | 86.4 (84.2 - 88.6) | 0.97 (0.90 - 1.04) | 1.05 (0.98 - 1.13) | 1.01 (0.94 - 1.08) |
| Furniture makers and other craft woodworkers | 15,405 | 13,194 | 85.6 (85.1 - 86.2) | 84.3 (82.8 - 85.7) | 1.26 (1.20 - 1.31) | 1.21 (1.15 - 1.26) | 1.23 (1.17 - 1.29) |
| Florists | 11,255 | 9,942 | 88.3 (87.7 - 88.9) | 87.8 (86.0 - 89.5) | 0.99 (0.93 - 1.05) | 1.10 (1.04 - 1.16) | 1.18 (1.11 - 1.25) |
| Other skilled trades n.e.c. | 19,993 | 17,230 | 86.2 (85.7 - 86.7) | 85.0 (83.7 - 86.3) | 1.20 (1.16 - 1.25) | 1.19 (1.14 - 1.24) | 1.22 (1.17 - 1.27) |
| Nursery nurses and assistants | 94,191 | 85,183 | 90.4 (90.2 - 90.6) | 90.8 (90.2 - 91.4) | 0.79 (0.77 - 0.81) | 0.81 (0.79 - 0.83) | 0.90 (0.88 - 0.92) |
| Childminders and related occupations | 61,026 | 53,818 | 88.2 (87.9 - 88.4) | 88.5 (87.7 - 89.3) | 1.00 (0.98 - 1.03) | 1.04 (1.01 - 1.06) | 0.98 (0.95 - 1.00) |
| Playworkers | 17,847 | 15,983 | 89.6 (89.1 - 90.0) | 89.2 (87.8 - 90.6) | 0.87 (0.83 - 0.92) | 0.97 (0.92 - 1.01) | 1.03 (0.98 - 1.09) |
| Teaching assistants | 212,606 | 195,875 | 92.1 (92.0 - 92.2) | 90.7 (90.3 - 91.1) | 0.64 (0.63 - 0.65) | 0.77 (0.76 - 0.79) | 0.87 (0.86 - 0.88) |
| Educational support assistants | 57,712 | 52,794 | 91.5 (91.3 - 91.7) | 90.4 (89.6 - 91.2) | 0.70 (0.68 - 0.72) | 0.83 (0.80 - 0.85) | 0.93 (0.90 - 0.95) |
| Veterinary nurses | 5,557 | 5,065 | 91.1 (90.4 - 91.9) | 92.5 (89.5 - 95.5) | 0.73 (0.66 - 0.80) | 0.61 (0.56 - 0.67) | 0.91 (0.83 - 1.00) |
| Pest control officers | 2,799 | 2,444 | 87.3 (86.1 - 88.5) | 85.5 (82.0 - 89.1) | 1.09 (0.97 - 1.22) | 1.06 (0.95 - 1.19) | 1.17 (1.04 - 1.31) |
| Animal care services occupations n.e.c. | 21,249 | 18,318 | 86.2 (85.7 - 86.7) | 86.3 (85.1 - 87.6) | 1.20 (1.15 - 1.25) | 1.21 (1.17 - 1.26) | 1.38 (1.33 - 1.44) |
| Nursing auxiliaries and assistants | 115,219 | 103,029 | 89.4 (89.2 - 89.6) | 88.8 (88.3 - 89.4) | 0.89 (0.87 - 0.90) | 0.99 (0.97 - 1.01) | 0.91 (0.89 - 0.93) |
| Ambulance staff (excluding paramedics) | 10,417 | 9,457 | 90.8 (90.2 - 91.3) | 90.6 (88.8 - 92.5) | 0.76 (0.71 - 0.81) | 0.76 (0.71 - 0.81) | 0.91 (0.85 - 0.97) |
| Dental nurses | 22,072 | 19,605 | 88.8 (88.4 - 89.2) | 89.7 (88.4 - 91.0) | 0.94 (0.90 - 0.98) | 0.92 (0.88 - 0.96) | 1.00 (0.96 - 1.05) |
| Houseparents and residential wardens | 22,051 | 19,750 | 89.6 (89.2 - 90.0) | 88.2 (86.9 - 89.6) | 0.87 (0.84 - 0.91) | 1.11 (1.06 - 1.16) | 1.14 (1.09 - 1.19) |
| Care workers and home carers | 395,569 | 339,216 | 85.8 (85.6 - 85.9) | 85.2 (84.9 - 85.5) | 1.25 (1.24 - 1.27) | 1.40 (1.39 - 1.41) | 1.21 (1.20 - 1.22) |
| Senior care workers | 32,461 | 29,235 | 90.1 (89.7 - 90.4) | 89.9 (88.8 - 90.9) | 0.83 (0.80 - 0.86) | 0.89 (0.86 - 0.92) | 0.92 (0.89 - 0.96) |
| Care escorts | 6,466 | 5,730 | 88.6 (87.8 - 89.4) | 85.5 (82.9 - 88.2) | 0.96 (0.89 - 1.04) | 1.31 (1.21 - 1.42) | 1.01 (0.93 - 1.09) |
| Undertakers, mortuary and crematorium assistants | 7,034 | 6,484 | 92.2 (91.6 - 92.8) | 91.3 (88.9 - 93.6) | 0.64 (0.58 - 0.69) | 0.69 (0.63 - 0.76) | 0.80 (0.73 - 0.87) |
| Sports and leisure assistants | 13,922 | 11,508 | 82.7 (82.0 - 83.3) | 83.5 (81.9 - 85.0) | 1.57 (1.51 - 1.64) | 1.42 (1.36 - 1.49) | 1.36 (1.30 - 1.42) |
| Travel agents | 25,046 | 22,329 | 89.2 (88.8 - 89.5) | 89.8 (88.6 - 91.0) | 0.91 (0.88 - 0.95) | 0.85 (0.82 - 0.89) | 0.83 (0.80 - 0.87) |
| Air travel assistants | 23,608 | 20,647 | 87.5 (87.0 - 87.9) | 87.6 (86.4 - 88.8) | 1.08 (1.03 - 1.12) | 1.08 (1.03 - 1.12) | 1.00 (0.96 - 1.04) |
| Rail travel assistants | 8,276 | 7,340 | 88.7 (88.0 - 89.4) | 88.0 (86.0 - 90.1) | 0.96 (0.89 - 1.02) | 0.93 (0.86 - 0.99) | 0.80 (0.74 - 0.85) |
| Leisure and travel service occupations n.e.c. | 6,249 | 5,266 | 84.3 (83.4 - 85.2) | 83.0 (80.7 - 85.3) | 1.40 (1.31 - 1.50) | 1.47 (1.37 - 1.58) | 1.30 (1.21 - 1.40) |
| Hairdressers and barbers | 99,320 | 85,824 | 86.4 (86.2 - 86.6) | 86.6 (86.0 - 87.1) | 1.18 (1.16 - 1.20) | 1.22 (1.20 - 1.25) | 1.24 (1.22 - 1.27) |
| Beauticians and related occupations | 33,683 | 27,593 | 81.9 (81.5 - 82.3) | 83.3 (82.3 - 84.3) | 1.66 (1.61 - 1.70) | 1.59 (1.55 - 1.64) | 1.58 (1.54 - 1.63) |
| Housekeepers and related occupations | 32,014 | 27,574 | 86.1 (85.8 - 86.5) | 83.8 (82.8 - 84.9) | 1.21 (1.17 - 1.25) | 1.51 (1.46 - 1.56) | 1.03 (0.99 - 1.06) |
| Caretakers | 28,282 | 24,576 | 86.9 (86.5 - 87.3) | 83.0 (81.8 - 84.2) | 1.13 (1.09 - 1.17) | 1.28 (1.24 - 1.33) | 1.04 (1.00 - 1.08) |
| Cleaning and housekeeping managers and supervisors | 33,381 | 28,140 | 84.3 (83.9 - 84.7) | 82.7 (81.7 - 83.7) | 1.40 (1.36 - 1.44) | 1.58 (1.53 - 1.63) | 1.07 (1.04 - 1.10) |
| Sales and retail assistants | 514,651 | 444,584 | 86.4 (86.3 - 86.5) | 85.9 (85.7 - 86.2) | 1.19 (1.18 - 1.20) | 1.26 (1.25 - 1.27) | 1.09 (1.08 - 1.10) |
| Retail cashiers and check-out operators | 67,073 | 58,635 | 87.4 (87.2 - 87.7) | 86.8 (86.1 - 87.5) | 1.08 (1.05 - 1.10) | 1.18 (1.16 - 1.21) | 0.95 (0.92 - 0.97) |
| Telephone salespersons | 15,754 | 13,035 | 82.7 (82.2 - 83.3) | 84.0 (82.5 - 85.4) | 1.56 (1.50 - 1.63) | 1.47 (1.41 - 1.53) | 1.45 (1.39 - 1.52) |
| Pharmacy and other dispensing assistants | 33,177 | 30,170 | 90.9 (90.6 - 91.2) | 90.2 (89.1 - 91.2) | 0.75 (0.72 - 0.78) | 0.85 (0.82 - 0.88) | 0.88 (0.85 - 0.91) |
| Vehicle and parts salespersons and advisers | 19,651 | 17,167 | 87.4 (86.9 - 87.8) | 87.5 (86.2 - 88.8) | 1.08 (1.04 - 1.13) | 0.93 (0.89 - 0.97) | 1.00 (0.96 - 1.05) |
| Collector salespersons and credit agents | 5,095 | 4,220 | 82.8 (81.8 - 83.9) | 83.2 (80.6 - 85.7) | 1.55 (1.45 - 1.67) | 1.47 (1.36 - 1.58) | 1.53 (1.41 - 1.65) |
| Debt, rent and other cash collectors | 14,444 | 12,631 | 87.4 (86.9 - 88.0) | 86.7 (85.2 - 88.2) | 1.08 (1.02 - 1.13) | 1.10 (1.05 - 1.16) | 1.06 (1.01 - 1.12) |
| Roundspersons and van salespersons | 5,611 | 4,845 | 86.3 (85.4 - 87.2) | 83.4 (80.8 - 85.9) | 1.19 (1.10 - 1.28) | 1.24 (1.15 - 1.34) | 1.09 (1.00 - 1.18) |
| Market and street traders and assistants | 8,831 | 7,084 | 80.2 (79.4 - 81.0) | 77.5 (75.6 - 79.4) | 1.85 (1.76 - 1.95) | 2.03 (1.92 - 2.14) | 1.45 (1.37 - 1.53) |
| Merchandisers and window dressers | 15,814 | 14,075 | 89.0 (88.5 - 89.5) | 89.2 (87.7 - 90.7) | 0.93 (0.88 - 0.97) | 0.91 (0.87 - 0.96) | 0.96 (0.91 - 1.01) |
| Sales related occupations n.e.c. | 17,556 | 14,924 | 85.0 (84.5 - 85.5) | 85.1 (83.7 - 86.4) | 1.32 (1.27 - 1.38) | 1.26 (1.21 - 1.32) | 1.27 (1.21 - 1.32) |
| Sales supervisors | 63,831 | 57,378 | 89.9 (89.7 - 90.1) | 89.9 (89.2 - 90.6) | 0.84 (0.82 - 0.86) | 0.82 (0.80 - 0.84) | 0.83 (0.80 - 0.85) |
| Call and contact centre occupations | 20,478 | 17,680 | 86.3 (85.9 - 86.8) | 87.3 (86.0 - 88.6) | 1.19 (1.14 - 1.23) | 1.10 (1.05 - 1.14) | 1.08 (1.04 - 1.13) |
| Telephonists | 9,095 | 7,702 | 84.7 (83.9 - 85.4) | 83.3 (81.4 - 85.2) | 1.36 (1.28 - 1.44) | 1.57 (1.48 - 1.67) | 1.31 (1.23 - 1.39) |
| Communication operators | 16,722 | 15,056 | 90.0 (89.6 - 90.5) | 90.1 (88.7 - 91.5) | 0.83 (0.79 - 0.87) | 0.81 (0.77 - 0.85) | 0.84 (0.80 - 0.89) |
| Market research interviewers | 5,289 | 4,404 | 83.3 (82.3 - 84.3) | 83.0 (80.5 - 85.4) | 1.51 (1.40 - 1.62) | 1.54 (1.43 - 1.66) | 1.36 (1.26 - 1.47) |
| Customer service occupations n.e.c. | 114,720 | 99,432 | 86.7 (86.5 - 86.9) | 87.6 (87.0 - 88.1) | 1.15 (1.13 - 1.17) | 1.07 (1.06 - 1.09) | 0.99 (0.97 - 1.00) |
| Customer service managers and supervisors | 40,451 | 37,202 | 92.0 (91.7 - 92.2) | 92.1 (91.2 - 93.1) | 0.65 (0.63 - 0.68) | 0.62 (0.60 - 0.65) | 0.70 (0.67 - 0.72) |
| Food, drink and tobacco process operatives | 90,746 | 73,959 | 81.5 (81.2 - 81.8) | 80.8 (80.3 - 81.4) | 1.71 (1.68 - 1.74) | 1.67 (1.64 - 1.70) | 1.17 (1.15 - 1.19) |
| Glass and ceramics process operatives | 4,809 | 4,161 | 86.5 (85.6 - 87.5) | 84.8 (82.1 - 87.4) | 1.17 (1.07 - 1.27) | 1.12 (1.03 - 1.22) | 0.99 (0.91 - 1.08) |
| Textile process operatives | 11,841 | 10,138 | 85.6 (85.0 - 86.2) | 83.6 (81.9 - 85.3) | 1.26 (1.20 - 1.33) | 1.39 (1.32 - 1.47) | 0.98 (0.93 - 1.03) |
| Chemical and related process operatives | 20,422 | 18,267 | 89.4 (89.0 - 89.9) | 87.6 (86.2 - 88.9) | 0.88 (0.85 - 0.92) | 0.90 (0.86 - 0.94) | 0.92 (0.88 - 0.97) |
| Rubber process operatives | 3,807 | 3,328 | 87.4 (86.4 - 88.5) | 85.6 (82.5 - 88.6) | 1.08 (0.98 - 1.19) | 1.08 (0.98 - 1.19) | 0.95 (0.86 - 1.05) |
| Plastics process operatives | 21,083 | 18,092 | 85.8 (85.3 - 86.3) | 84.6 (83.3 - 85.8) | 1.24 (1.19 - 1.29) | 1.19 (1.14 - 1.24) | 1.04 (1.00 - 1.09) |
| Metal making and treating process operatives | 6,564 | 5,835 | 88.9 (88.1 - 89.7) | 86.4 (84.0 - 88.8) | 0.94 (0.87 - 1.01) | 0.96 (0.89 - 1.04) | 0.93 (0.86 - 1.01) |
| Electroplaters | 2,880 | 2,407 | 83.6 (82.2 - 84.9) | 82.4 (79.1 - 85.8) | 1.47 (1.33 - 1.63) | 1.35 (1.22 - 1.50) | 1.22 (1.10 - 1.35) |
| Process operatives n.e.c. | 3,921 | 3,456 | 88.1 (87.1 - 89.2) | 86.7 (83.7 - 89.7) | 1.01 (0.92 - 1.11) | 0.98 (0.89 - 1.08) | 0.97 (0.88 - 1.07) |
| Paper and wood machine operatives | 17,221 | 15,062 | 87.5 (87.0 - 88.0) | 85.6 (84.2 - 87.1) | 1.07 (1.03 - 1.12) | 1.05 (1.01 - 1.10) | 0.98 (0.93 - 1.03) |
| Coal mine operatives | 3,431 | 3,139 | 91.5 (90.6 - 92.4) | 86.0 (80.2 - 91.9) | 0.70 (0.62 - 0.79) | 0.92 (0.82 - 1.04) | 0.75 (0.66 - 0.84) |
| Quarry workers and related operatives | 4,941 | 4,332 | 87.7 (86.8 - 88.6) | 86.2 (83.6 - 88.8) | 1.05 (0.97 - 1.15) | 0.99 (0.91 - 1.08) | 1.08 (0.99 - 1.17) |
| Energy plant operatives | 3,360 | 3,048 | 90.7 (89.7 - 91.7) | 89.2 (85.9 - 92.6) | 0.77 (0.68 - 0.86) | 0.77 (0.68 - 0.86) | 0.88 (0.79 - 1.00) |
| Metal working machine operatives | 73,019 | 63,149 | 86.5 (86.2 - 86.7) | 84.9 (84.2 - 85.6) | 1.17 (1.15 - 1.20) | 1.13 (1.11 - 1.15) | 1.12 (1.09 - 1.14) |
| Water and sewerage plant operatives | 6,252 | 5,631 | 90.1 (89.3 - 90.8) | 89.1 (86.7 - 91.4) | 0.83 (0.76 - 0.90) | 0.78 (0.72 - 0.85) | 0.91 (0.83 - 0.99) |
| Printing machine assistants | 7,752 | 6,718 | 86.7 (85.9 - 87.4) | 85.8 (83.7 - 87.8) | 1.15 (1.08 - 1.23) | 1.12 (1.05 - 1.20) | 1.01 (0.94 - 1.08) |
| Plant and machine operatives n.e.c. | 25,214 | 21,205 | 84.1 (83.6 - 84.6) | 82.6 (81.5 - 83.7) | 1.42 (1.37 - 1.47) | 1.39 (1.34 - 1.44) | 1.09 (1.05 - 1.13) |
| Assemblers (electrical and electronic products) | 17,682 | 15,456 | 87.4 (86.9 - 87.9) | 85.7 (84.3 - 87.2) | 1.08 (1.03 - 1.13) | 1.17 (1.12 - 1.23) | 1.06 (1.01 - 1.11) |
| Assemblers (vehicles and metal goods) | 21,716 | 19,072 | 87.8 (87.4 - 88.3) | 85.9 (84.7 - 87.2) | 1.04 (1.00 - 1.08) | 1.03 (0.99 - 1.08) | 0.93 (0.89 - 0.97) |
| Routine inspectors and testers | 29,716 | 26,188 | 88.1 (87.8 - 88.5) | 86.6 (85.6 - 87.7) | 1.01 (0.97 - 1.05) | 1.03 (1.00 - 1.07) | 1.01 (0.97 - 1.04) |
| Weighers, graders and sorters | 4,002 | 3,313 | 82.8 (81.6 - 84.0) | 81.3 (78.5 - 84.1) | 1.56 (1.44 - 1.69) | 1.64 (1.51 - 1.78) | 1.27 (1.17 - 1.39) |
| Tyre, exhaust and windscreen fitters | 6,943 | 5,692 | 82.0 (81.1 - 82.9) | 82.9 (80.7 - 85.1) | 1.65 (1.55 - 1.75) | 1.35 (1.26 - 1.43) | 1.32 (1.23 - 1.40) |
| Sewing machinists | 31,825 | 27,250 | 85.6 (85.2 - 86.0) | 83.3 (82.2 - 84.3) | 1.26 (1.22 - 1.30) | 1.58 (1.53 - 1.63) | 1.02 (0.98 - 1.05) |
| Assemblers and routine operatives n.e.c. | 23,201 | 19,599 | 84.5 (84.0 - 84.9) | 83.1 (81.9 - 84.3) | 1.38 (1.33 - 1.43) | 1.40 (1.36 - 1.46) | 1.16 (1.11 - 1.20) |
| Scaffolders, staggers and riggers | 14,606 | 11,737 | 80.4 (79.7 - 81.0) | 80.8 (79.4 - 82.3) | 1.83 (1.76 - 1.91) | 1.54 (1.48 - 1.61) | 1.61 (1.54 - 1.68) |
| Road construction operatives | 12,119 | 10,309 | 85.1 (84.4 - 85.7) | 83.6 (81.9 - 85.2) | 1.32 (1.25 - 1.38) | 1.22 (1.16 - 1.28) | 1.21 (1.15 - 1.27) |
| Rail construction and maintenance operatives | 4,310 | 3,688 | 85.6 (84.5 - 86.6) | 84.4 (81.6 - 87.1) | 1.26 (1.16 - 1.38) | 1.18 (1.08 - 1.28) | 1.00 (0.92 - 1.10) |
| Construction operatives n.e.c. | 54,885 | 45,416 | 82.7 (82.4 - 83.1) | 81.0 (80.3 - 81.8) | 1.57 (1.53 - 1.60) | 1.48 (1.45 - 1.51) | 1.33 (1.30 - 1.36) |
| Large goods vehicle drivers | 133,780 | 118,999 | 89.0 (88.8 - 89.1) | 86.1 (85.6 - 86.7) | 0.93 (0.91 - 0.95) | 0.96 (0.94 - 0.97) | 0.95 (0.93 - 0.97) |
| Van drivers | 141,151 | 118,952 | 84.3 (84.1 - 84.5) | 82.3 (81.9 - 82.8) | 1.40 (1.38 - 1.42) | 1.38 (1.36 - 1.40) | 1.13 (1.11 - 1.14) |
| Bus and coach drivers | 67,863 | 56,628 | 83.4 (83.2 - 83.7) | 81.0 (80.3 - 81.7) | 1.49 (1.46 - 1.52) | 1.52 (1.49 - 1.55) | 1.07 (1.05 - 1.09) |
| Taxi and cab drivers and chauffeurs | 100,645 | 83,874 | 83.3 (83.1 - 83.6) | 81.5 (80.9 - 82.1) | 1.50 (1.48 - 1.53) | 1.46 (1.43 - 1.48) | 0.88 (0.86 - 0.90) |
| Driving instructors | 18,123 | 16,307 | 90.0 (89.5 - 90.4) | 88.2 (86.7 - 89.6) | 0.83 (0.80 - 0.88) | 0.89 (0.85 - 0.93) | 0.99 (0.94 - 1.04) |
| Crane drivers | 5,343 | 4,536 | 84.9 (83.9 - 85.9) | 83.1 (80.6 - 85.6) | 1.33 (1.24 - 1.44) | 1.28 (1.18 - 1.38) | 1.21 (1.12 - 1.31) |
| Fork-lift truck drivers | 39,486 | 32,148 | 81.4 (81.0 - 81.8) | 80.5 (79.7 - 81.4) | 1.71 (1.67 - 1.76) | 1.54 (1.50 - 1.58) | 1.25 (1.22 - 1.29) |
| Agricultural machinery drivers | 1,155 | 1,031 | 89.3 (87.5 - 91.0) | 88.9 (83.3 - 94.4) | 0.90 (0.75 - 1.09) | 0.81 (0.67 - 0.98) | 0.83 (0.68 - 1.00) |
| Mobile machine drivers and operatives n.e.c. | 24,011 | 20,447 | 85.2 (84.7 - 85.6) | 83.1 (82.0 - 84.3) | 1.31 (1.26 - 1.35) | 1.26 (1.21 - 1.30) | 1.14 (1.10 - 1.18) |
| Train and tram drivers | 13,616 | 12,349 | 90.7 (90.2 - 91.2) | 89.0 (87.2 - 90.7) | 0.77 (0.73 - 0.81) | 0.80 (0.76 - 0.85) | 0.80 (0.75 - 0.85) |
| Marine and waterways transport operatives | 3,572 | 3,086 | 86.4 (85.3 - 87.5) | 84.6 (81.5 - 87.8) | 1.18 (1.07 - 1.30) | 1.19 (1.08 - 1.31) | 1.30 (1.18 - 1.44) |
| Air transport operatives | 6,823 | 5,999 | 87.9 (87.1 - 88.7) | 86.7 (84.5 - 89.0) | 1.03 (0.96 - 1.11) | 0.99 (0.92 - 1.07) | 0.82 (0.76 - 0.88) |
| Rail transport operatives | 10,122 | 8,943 | 88.4 (87.7 - 89.0) | 87.0 (85.1 - 88.9) | 0.99 (0.93 - 1.05) | 0.97 (0.91 - 1.03) | 0.93 (0.87 - 0.99) |
| Other drivers and transport operatives n.e.c. | 7,491 | 6,632 | 88.5 (87.8 - 89.3) | 87.3 (85.2 - 89.5) | 0.97 (0.90 - 1.04) | 0.94 (0.87 - 1.01) | 0.91 (0.84 - 0.98) |
| Farm workers | 17,701 | 15,667 | 88.5 (88.0 - 89.0) | 87.2 (85.8 - 88.6) | 0.97 (0.93 - 1.02) | 0.97 (0.92 - 1.01) | 0.95 (0.90 - 0.99) |
| Forestry workers | 3,885 | 3,246 | 83.6 (82.4 - 84.7) | 83.2 (80.3 - 86.1) | 1.48 (1.36 - 1.61) | 1.31 (1.20 - 1.43) | 1.59 (1.45 - 1.73) |
| Fishing and other elementary agriculture occupations n.e.c. | 10,168 | 8,473 | 83.3 (82.6 - 84.1) | 81.7 (79.9 - 83.5) | 1.50 (1.42 - 1.58) | 1.53 (1.45 - 1.61) | 1.37 (1.30 - 1.45) |
| Elementary construction occupations | 80,244 | 63,166 | 78.7 (78.4 - 79.0) | 77.7 (77.1 - 78.4) | 2.04 (2.00 - 2.07) | 1.86 (1.83 - 1.89) | 1.52 (1.49 - 1.55) |
| Industrial cleaning process occupations | 10,974 | 8,862 | 80.8 (80.0 - 81.5) | 78.6 (76.9 - 80.3) | 1.79 (1.70 - 1.87) | 1.81 (1.72 - 1.90) | 1.40 (1.33 - 1.47) |
| Packers, bottlers, canners and fillers | 72,464 | 55,838 | 77.1 (76.8 - 77.4) | 76.5 (75.9 - 77.1) | 2.24 (2.21 - 2.28) | 2.33 (2.29 - 2.37) | 1.43 (1.40 - 1.45) |
| Elementary process plant occupations n.e.c. | 82,130 | 67,682 | 82.4 (82.1 - 82.7) | 81.2 (80.6 - 81.8) | 1.61 (1.58 - 1.63) | 1.59 (1.56 - 1.62) | 1.21 (1.19 - 1.23) |
| Postal workers, mail sorters, messengers and couriers | 105,441 | 91,721 | 87.0 (86.8 - 87.2) | 85.4 (84.8 - 86.0) | 1.12 (1.10 - 1.14) | 1.13 (1.11 - 1.15) | 1.00 (0.98 - 1.02) |
| Elementary administration occupations n.e.c. | 25,254 | 21,643 | 85.7 (85.3 - 86.1) | 84.7 (83.6 - 85.9) | 1.25 (1.21 - 1.30) | 1.35 (1.30 - 1.40) | 1.17 (1.13 - 1.21) |
| Window cleaners | 16,746 | 13,648 | 81.5 (80.9 - 82.1) | 80.0 (78.7 - 81.4) | 1.70 (1.64 - 1.77) | 1.60 (1.54 - 1.67) | 1.53 (1.47 - 1.60) |
| Street cleaners | 5,861 | 4,848 | 82.7 (81.7 - 83.7) | 79.3 (76.9 - 81.8) | 1.57 (1.46 - 1.68) | 1.65 (1.54 - 1.76) | 1.21 (1.12 - 1.30) |
| Cleaners and domestics | 330,820 | 273,741 | 82.7 (82.6 - 82.9) | 81.0 (80.7 - 81.3) | 1.58 (1.57 - 1.60) | 1.87 (1.85 - 1.88) | 1.23 (1.22 - 1.24) |
| Launderers, dry cleaners and pressers | 22,458 | 19,011 | 84.7 (84.2 - 85.1) | 82.8 (81.6 - 84.0) | 1.36 (1.31 - 1.41) | 1.56 (1.51 - 1.62) | 1.06 (1.02 - 1.11) |
| Refuse and salvage occupations | 19,589 | 16,071 | 82.0 (81.5 - 82.6) | 80.8 (79.5 - 82.0) | 1.64 (1.58 - 1.70) | 1.53 (1.48 - 1.59) | 1.31 (1.26 - 1.37) |
| Vehicle valeters and cleaners | 14,511 | 11,402 | 78.6 (77.9 - 79.2) | 79.2 (77.8 - 80.7) | 2.05 (1.97 - 2.13) | 1.76 (1.69 - 1.83) | 1.28 (1.23 - 1.33) |
| Elementary cleaning occupations n.e.c. | 1,158 | 997 | 86.1 (84.1 - 88.1) | 83.7 (78.2 - 89.2) | 1.21 (1.02 - 1.43) | 1.27 (1.07 - 1.50) | 1.16 (0.98 - 1.38) |
| Security guards and related occupations | 90,565 | 74,080 | 81.8 (81.5 - 82.0) | 81.0 (80.4 - 81.6) | 1.67 (1.65 - 1.70) | 1.57 (1.54 - 1.59) | 1.11 (1.09 - 1.13) |
| Parking and civil enforcement occupations | 7,600 | 6,370 | 83.8 (83.0 - 84.6) | 82.6 (80.6 - 84.7) | 1.45 (1.36 - 1.54) | 1.41 (1.33 - 1.50) | 1.05 (0.99 - 1.12) |
| School midday and crossing patrol occupations | 72,304 | 64,982 | 89.9 (89.7 - 90.1) | 88.6 (87.8 - 89.3) | 0.84 (0.82 - 0.86) | 1.02 (0.99 - 1.04) | 0.90 (0.88 - 0.93) |
| Elementary security occupations n.e.c. | 9,977 | 8,099 | 81.2 (80.4 - 81.9) | 80.6 (78.9 - 82.4) | 1.74 (1.65 - 1.83) | 1.66 (1.57 - 1.74) | 1.40 (1.32 - 1.47) |
| Shelf fillers | 37,846 | 32,812 | 86.7 (86.4 - 87.0) | 85.8 (84.9 - 86.8) | 1.15 (1.12 - 1.19) | 1.20 (1.16 - 1.23) | 1.04 (1.01 - 1.07) |
| Elementary sales occupations n.e.c. | 8,219 | 6,998 | 85.1 (84.4 - 85.9) | 84.3 (82.4 - 86.3) | 1.31 (1.23 - 1.39) | 1.32 (1.25 - 1.41) | 1.12 (1.05 - 1.19) |
| Elementary storage occupations | 216,101 | 178,936 | 82.8 (82.6 - 83.0) | 82.4 (82.1 - 82.8) | 1.57 (1.55 - 1.59) | 1.43 (1.41 - 1.44) | 1.16 (1.14 - 1.17) |
| Hospital porters | 6,275 | 5,523 | 88.0 (87.2 - 88.8) | 86.2 (83.7 - 88.6) | 1.02 (0.95 - 1.10) | 1.04 (0.97 - 1.13) | 0.81 (0.75 - 0.88) |
| Kitchen and catering assistants | 158,517 | 135,022 | 85.2 (85.0 - 85.4) | 84.3 (83.8 - 84.7) | 1.31 (1.29 - 1.33) | 1.43 (1.41 - 1.45) | 1.05 (1.04 - 1.07) |
| Waiters and waitresses | 52,218 | 41,224 | 78.9 (78.6 - 79.3) | 81.0 (80.2 - 81.9) | 2.01 (1.96 - 2.05) | 1.80 (1.76 - 1.84) | 1.18 (1.15 - 1.20) |
| Bar staff | 55,784 | 45,076 | 80.8 (80.5 - 81.1) | 82.5 (81.7 - 83.3) | 1.79 (1.75 - 1.82) | 1.66 (1.62 - 1.69) | 1.51 (1.47 - 1.54) |
| Leisure and theme park attendants | 4,797 | 3,881 | 80.9 (79.8 - 82.0) | 81.9 (79.3 - 84.5) | 1.77 (1.65 - 1.90) | 1.60 (1.49 - 1.72) | 1.39 (1.28 - 1.49) |
| Other elementary services occupations<br>n.e.c. | 10,745 | 8,792 | 81.8 (81.1 - 82.6) | 81.4 (79.7 - 83.1) | 1.67 (1.59 - 1.75) | 1.66 (1.58 - 1.75) | 1.33 (1.27 - 1.40) |

Note: ONS Public Health Data Asset; Vaccination rate is the rate of people fully vaccinated. The outcome of the logistic regression models is not being fully vaccinated as of 31 August 2021. The reference category for the ORs is all other occupations. Fully adjusted models include sex, five-year age bands, ethnicity, region, education, disability, pre-existing condition.

**Table S4:**
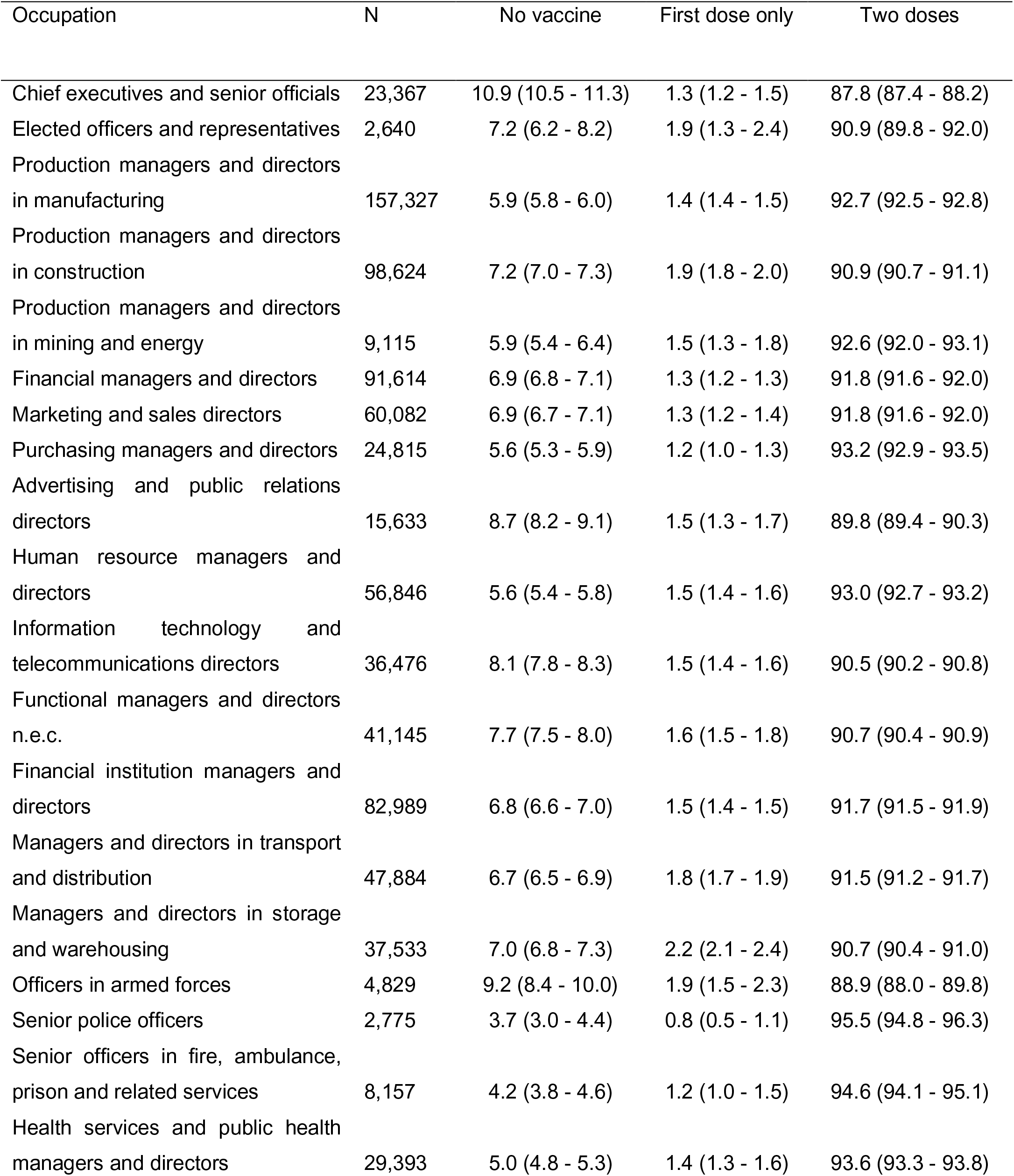
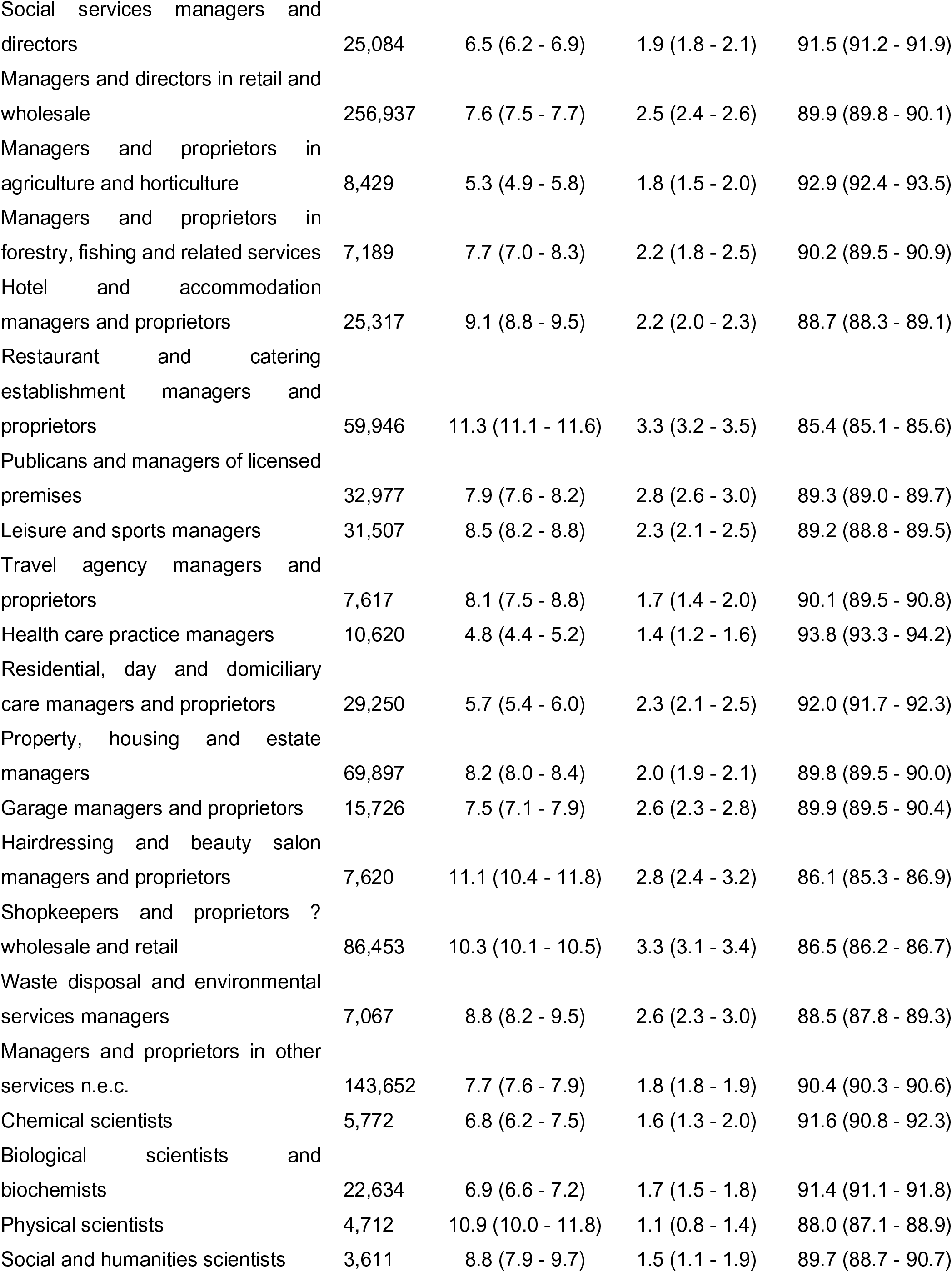
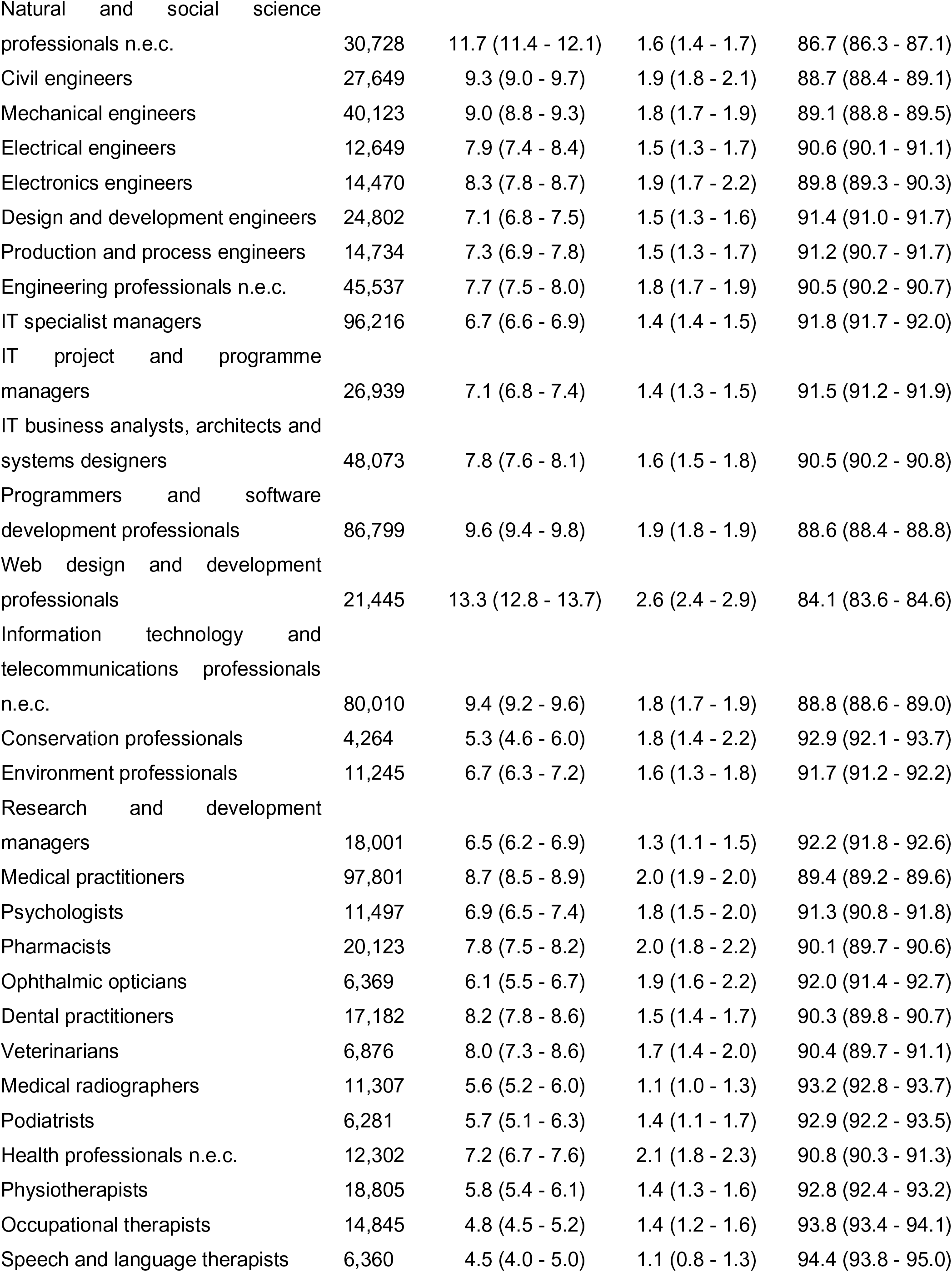
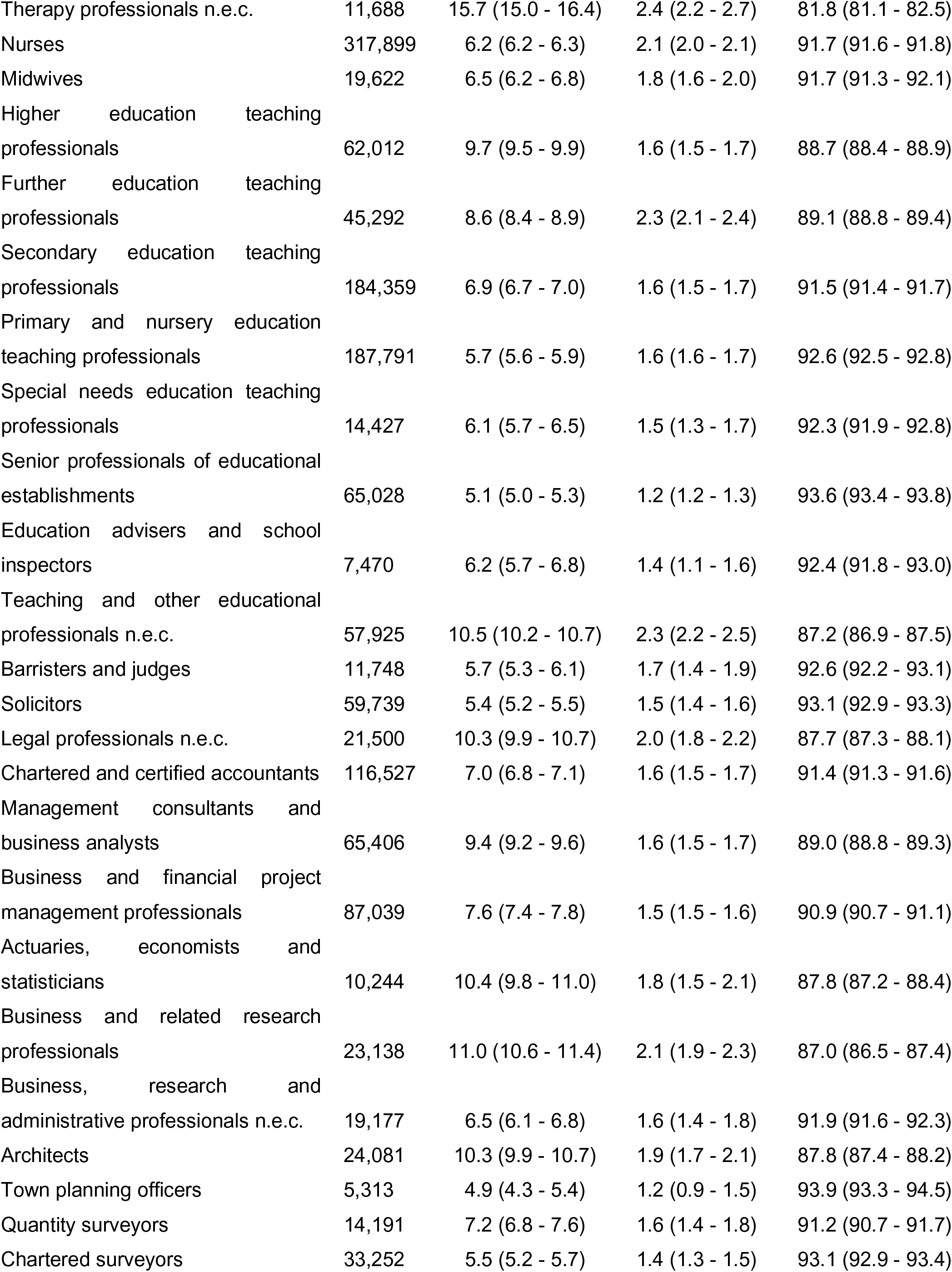
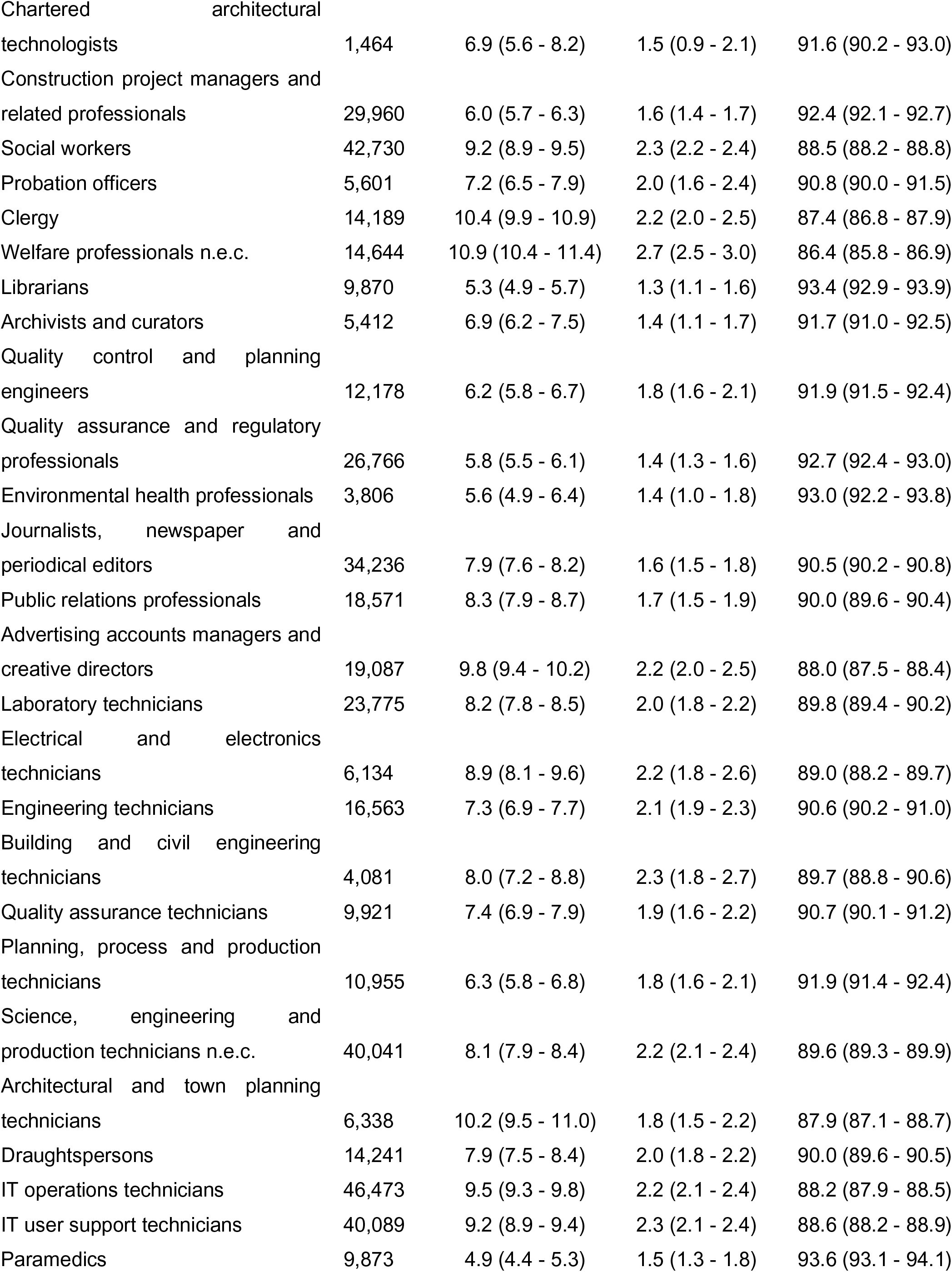
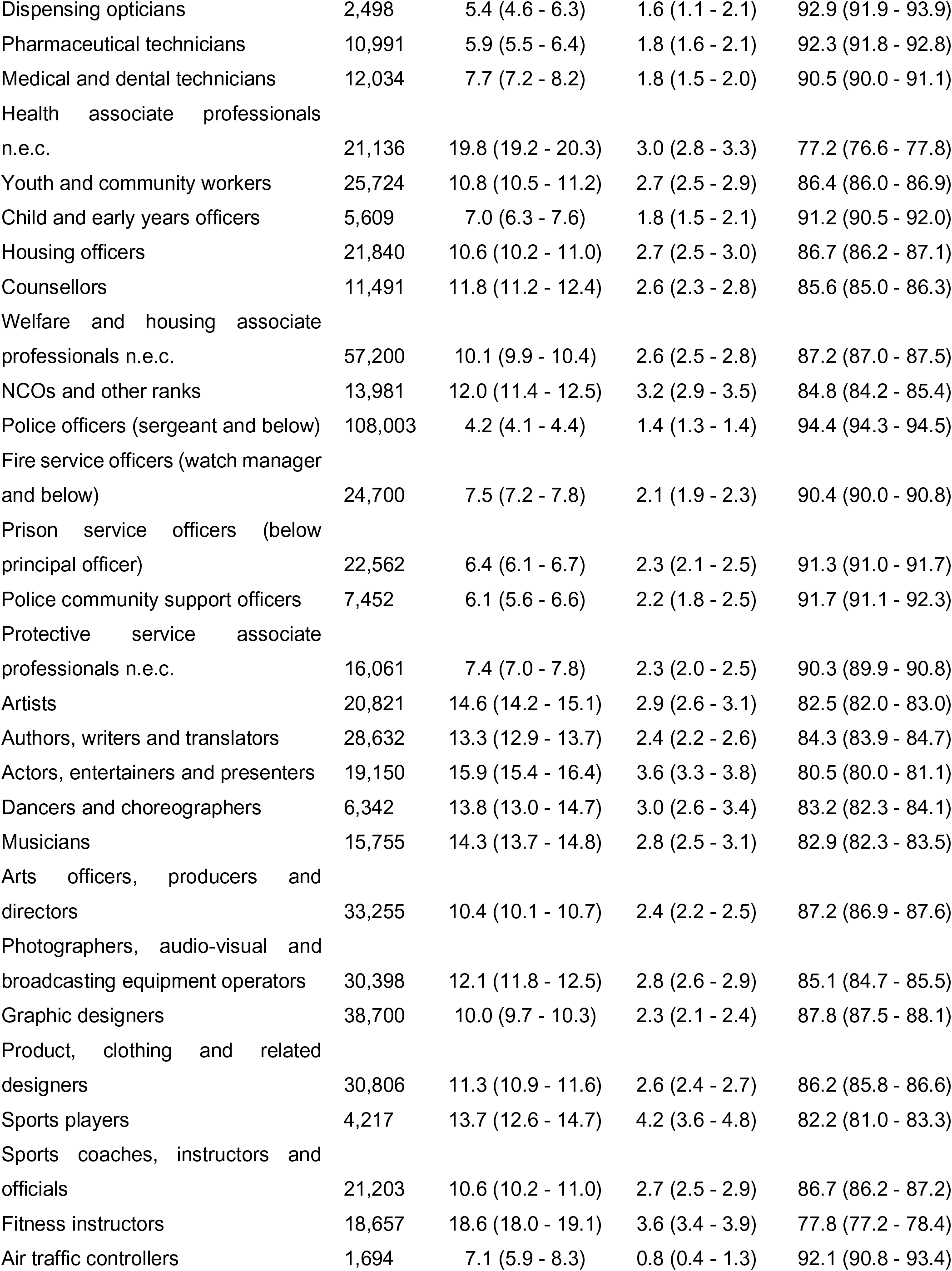
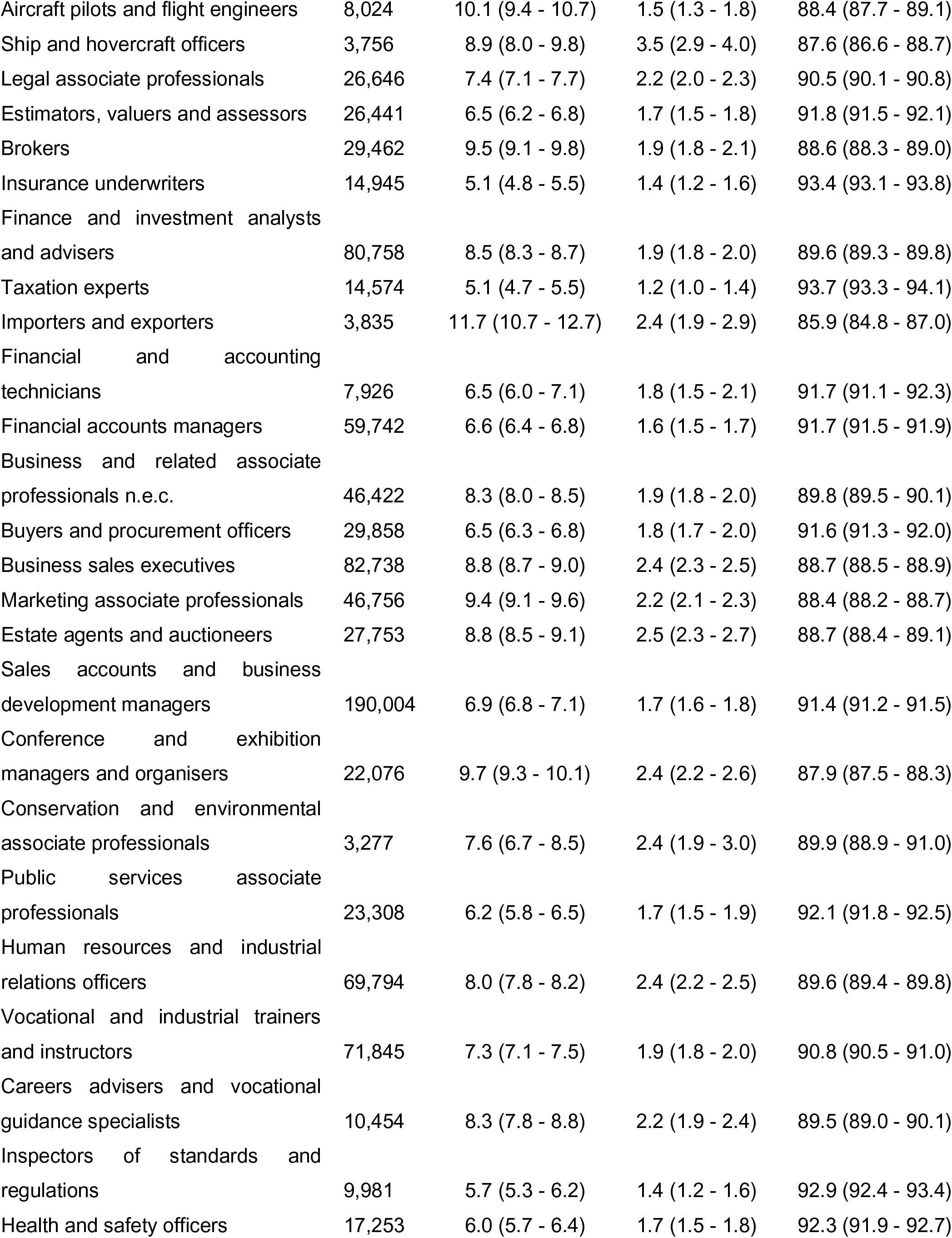
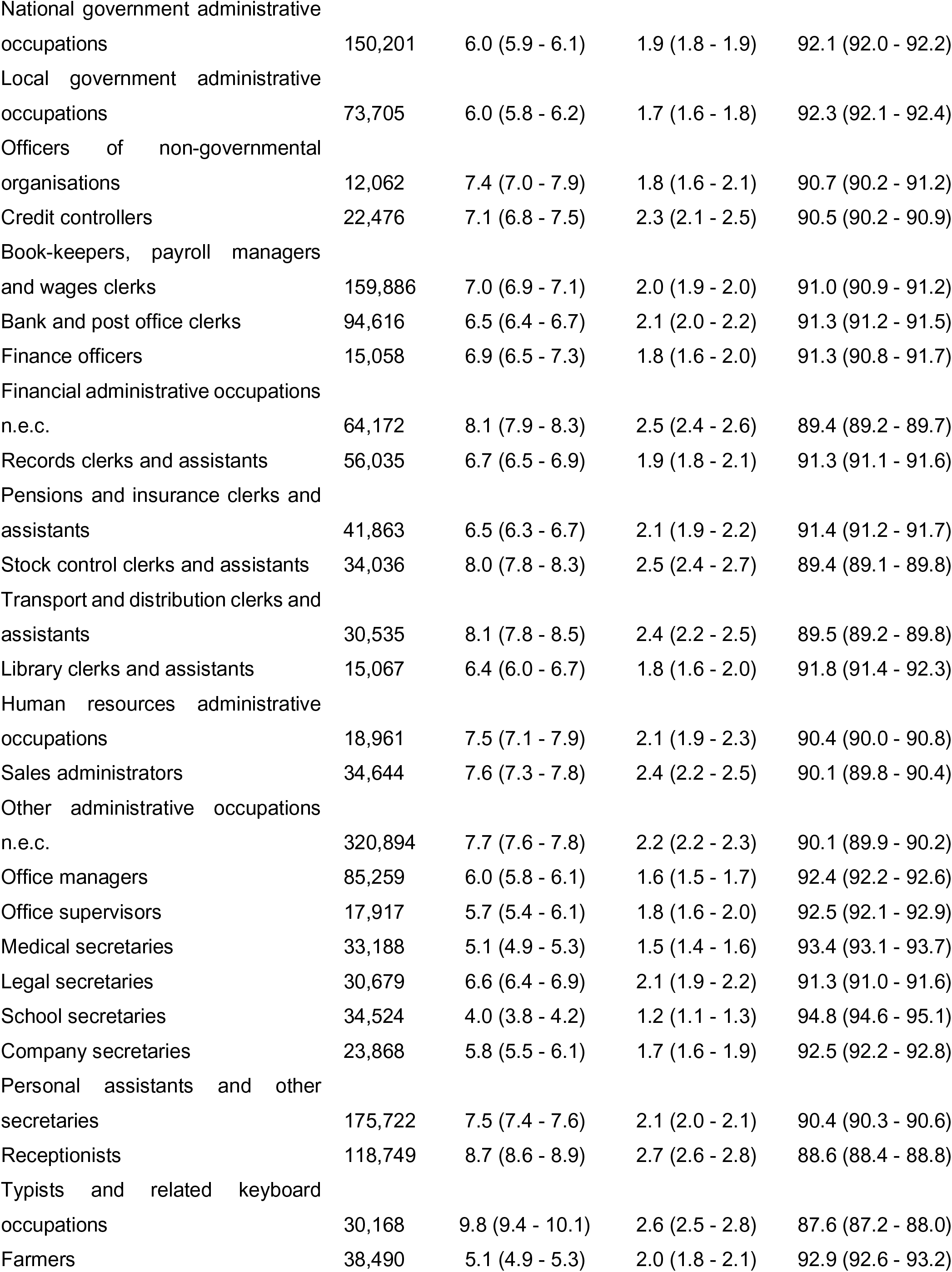
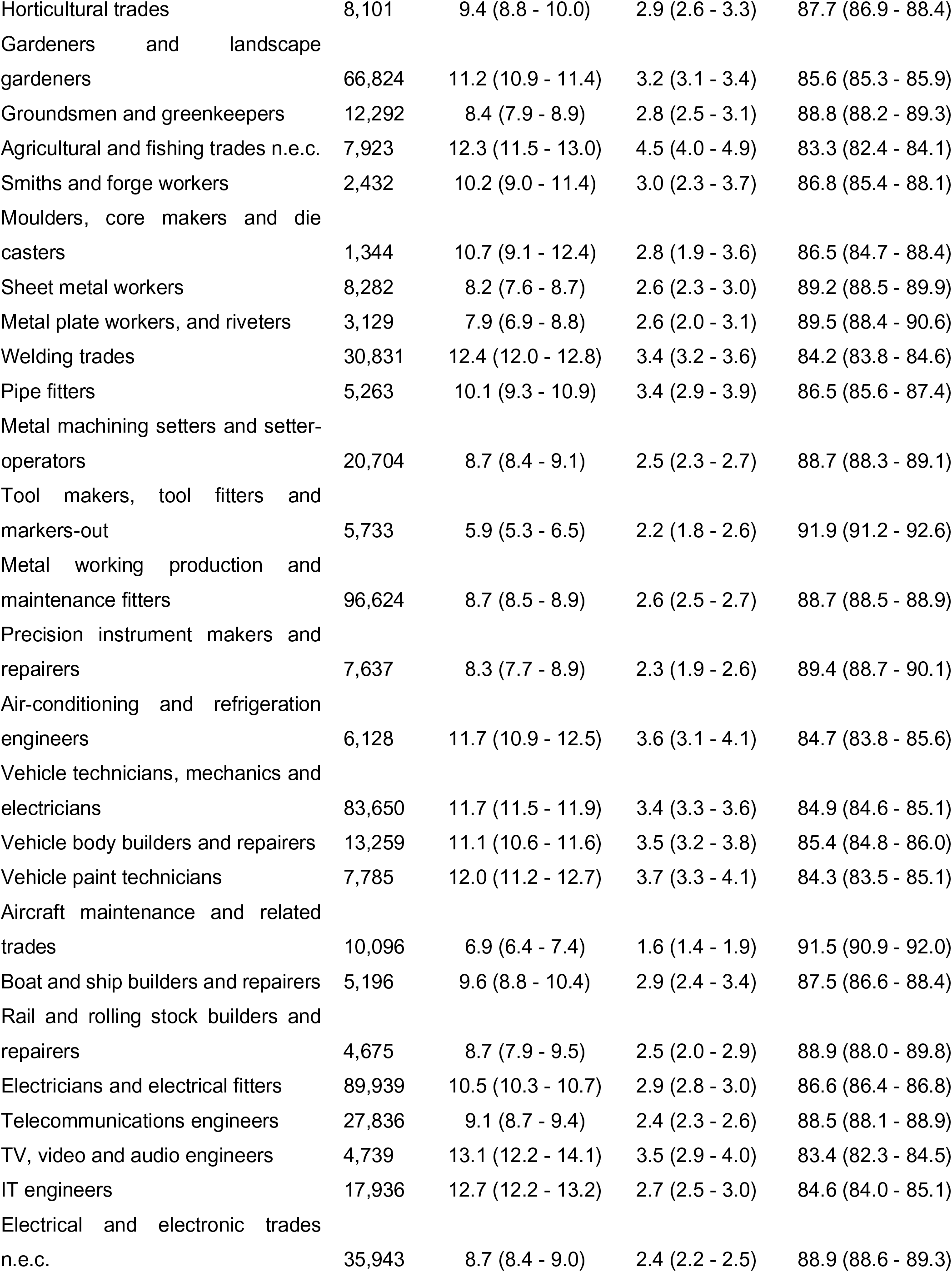
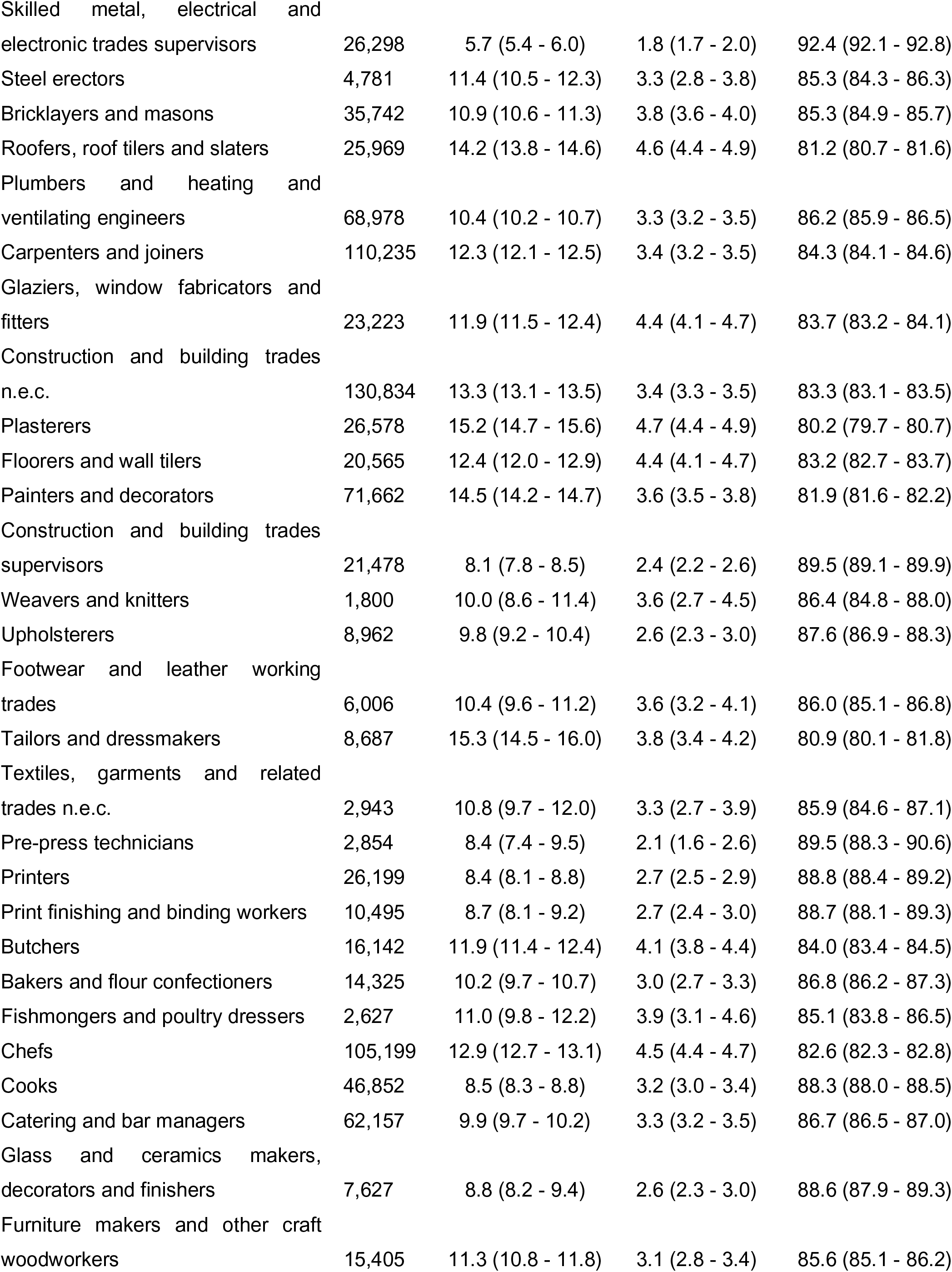
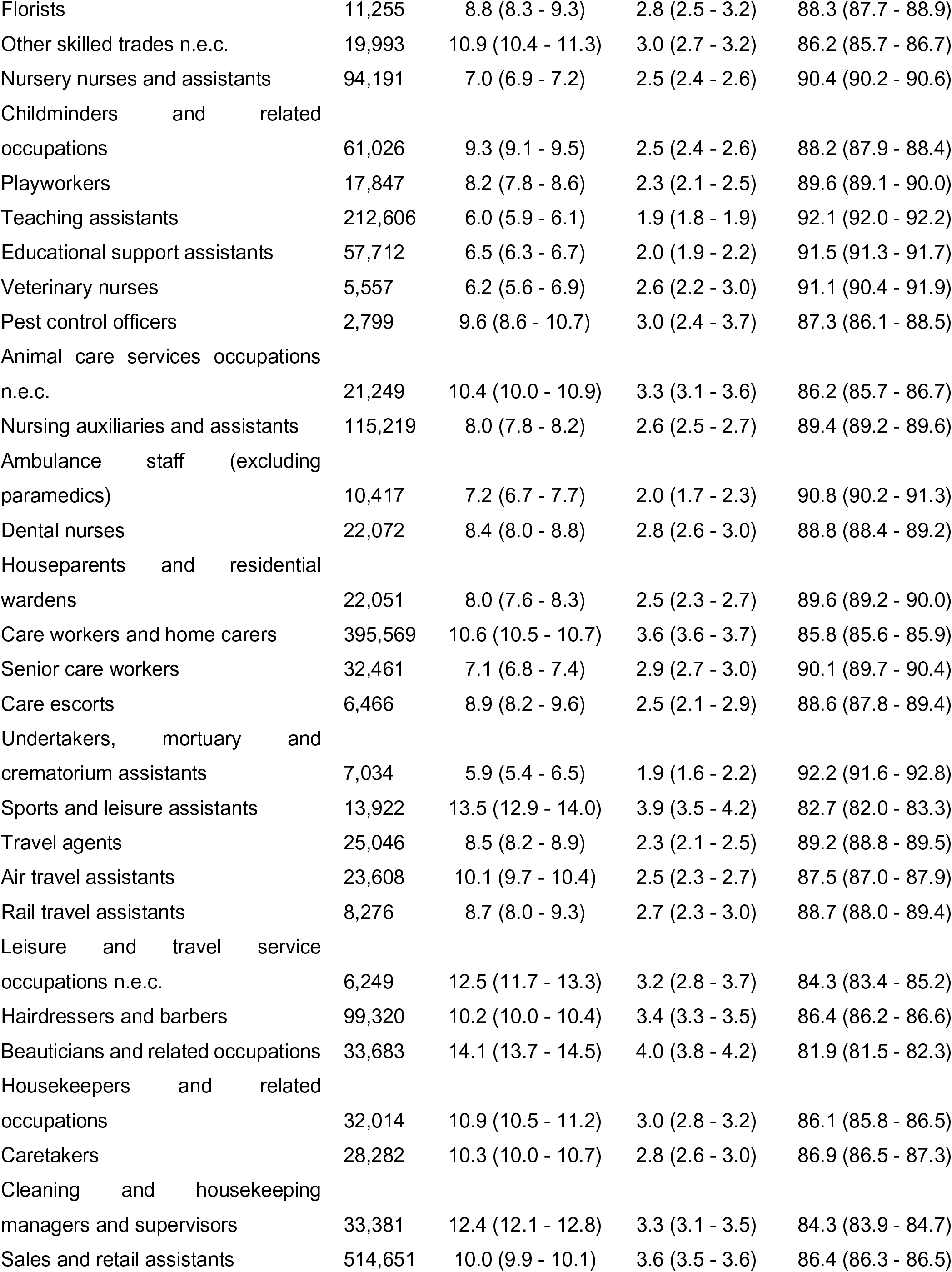
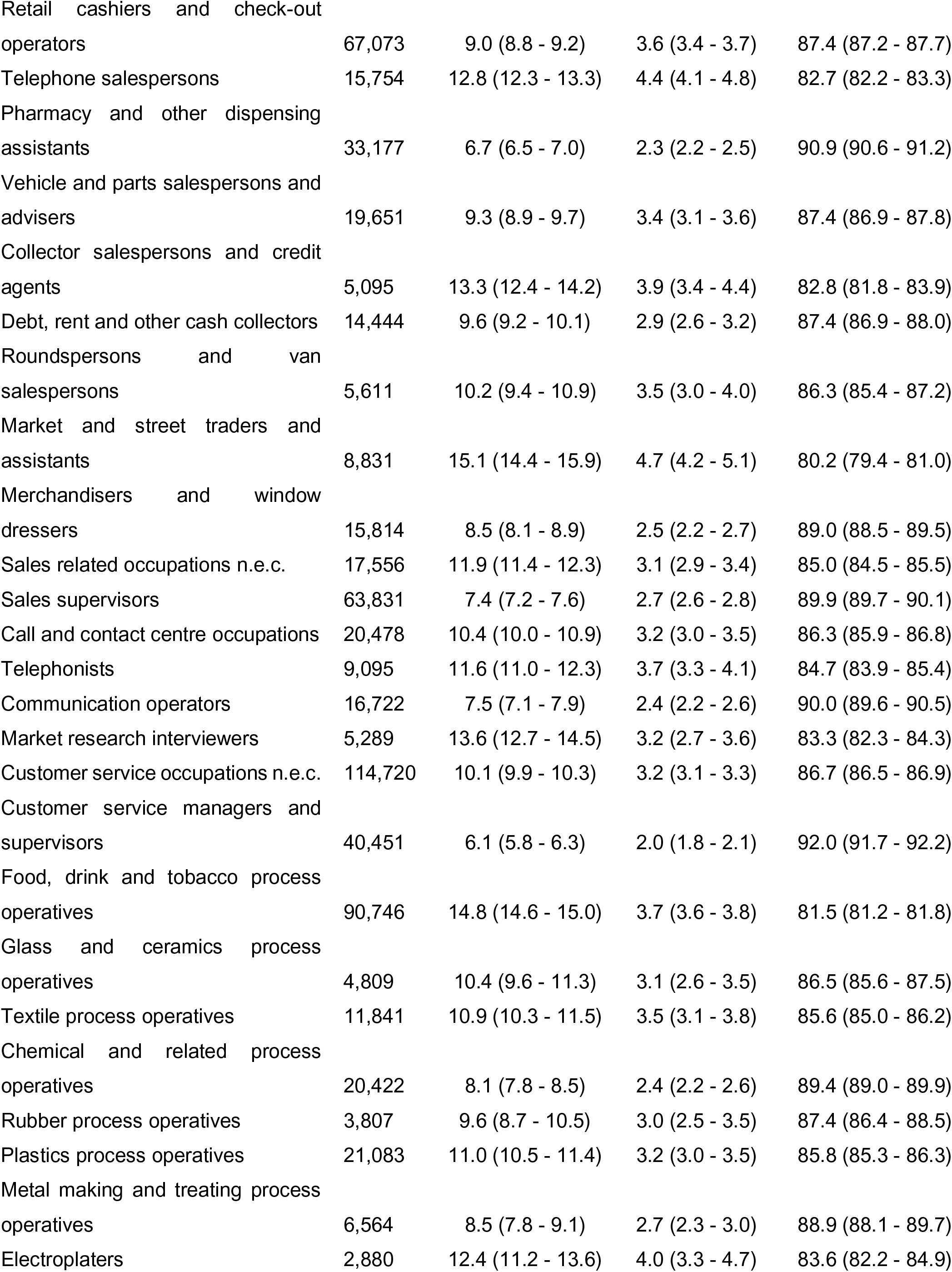
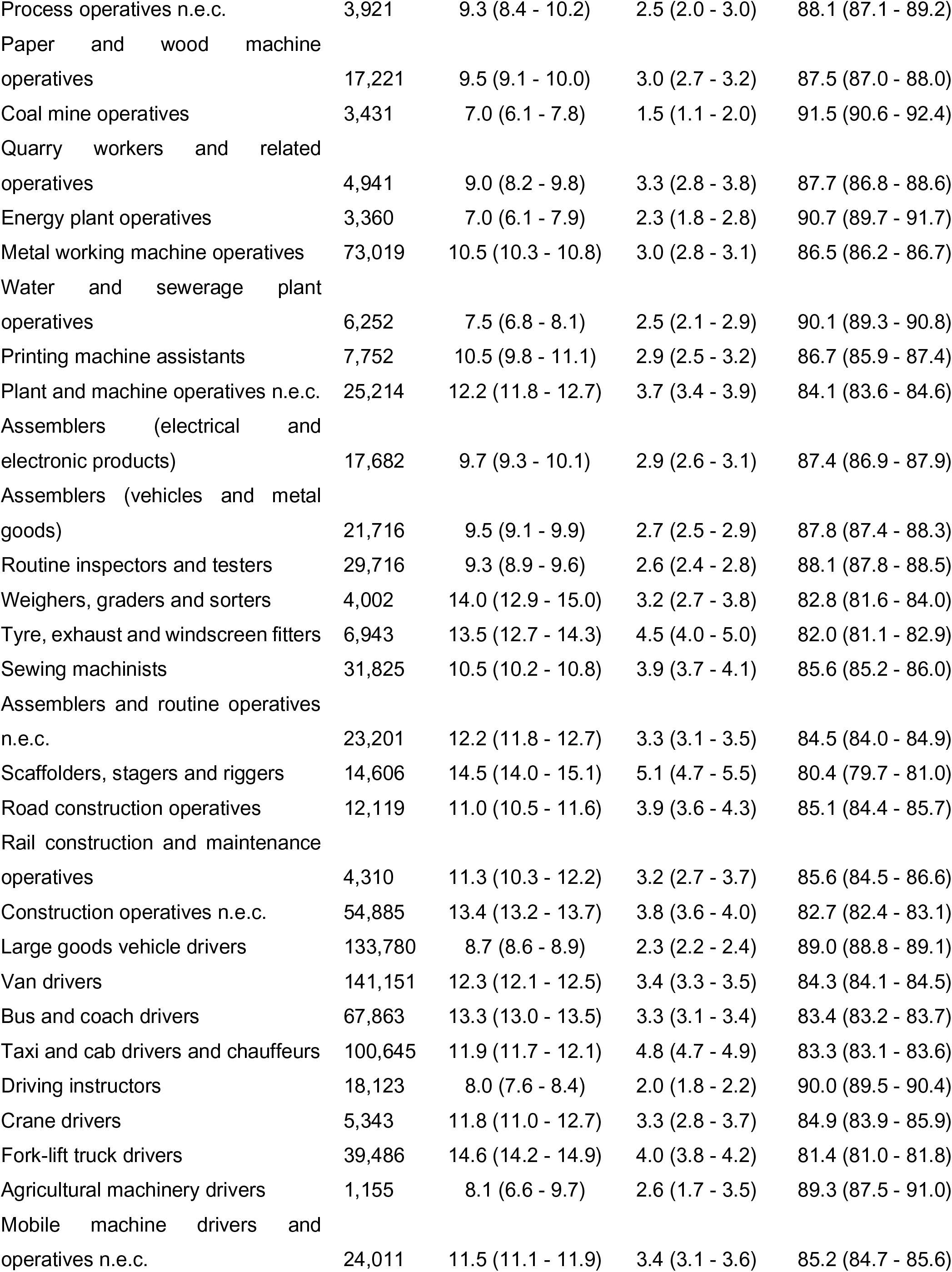
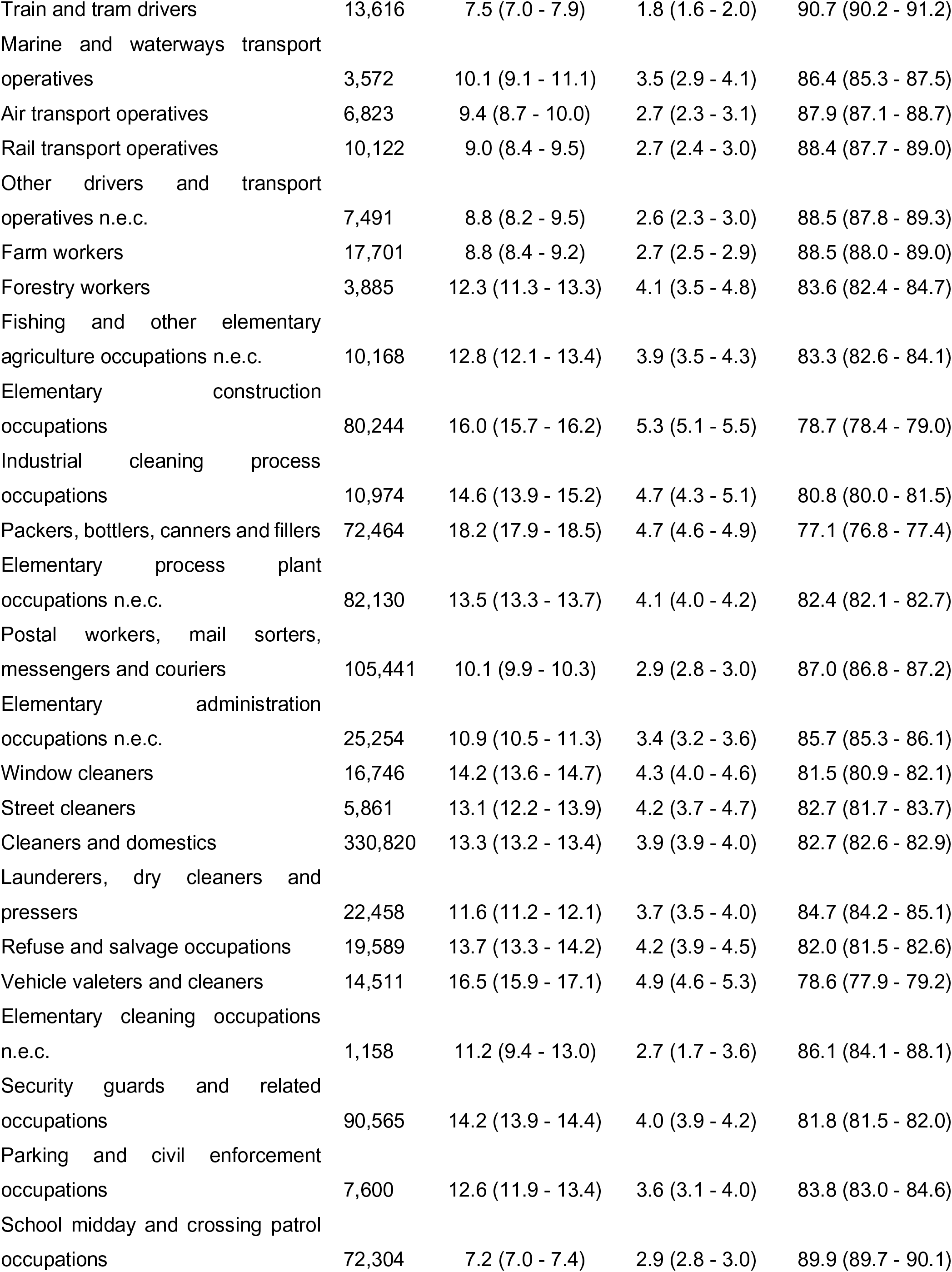
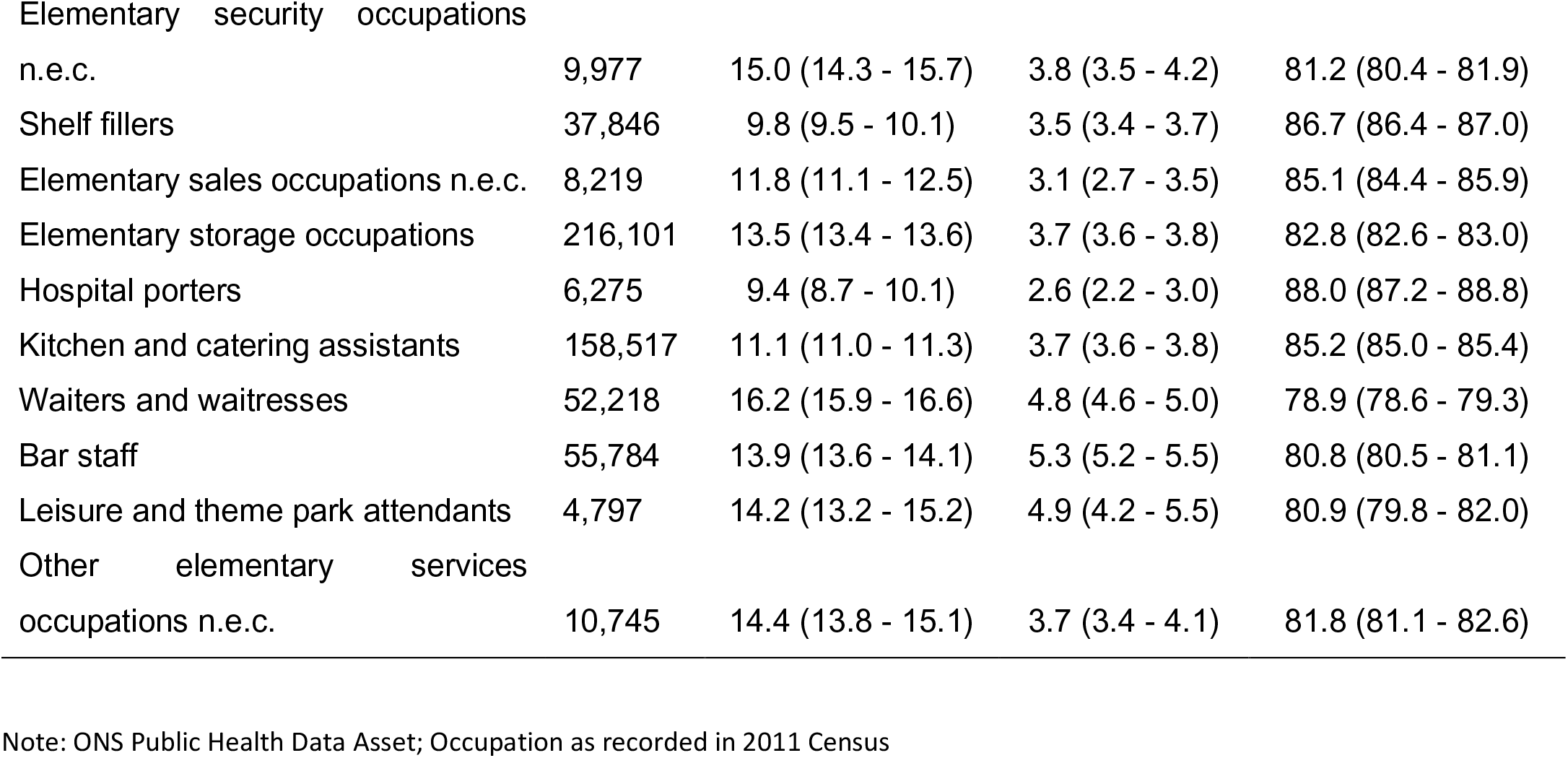
Vaccination status by SOC unit group.

